# A Biologically Constrained Continuous-Time Framework for Long-Horizon Cognition Forecasting in Alzheimer’s Disease

**DOI:** 10.64898/2026.08.07.26359964

**Authors:** Pon Deepika, Suresh Sunkari, Sai Koundinya Upadhyayula, the Alzheimer’s Disease Neuroimaging Initiative, Vaanathi Sundaresan

## Abstract

Accurate long-term forecasting of cognitive trajectories across the Alzheimer’s disease continuum is essential for early intervention, personalized prognosis, patient stratification, and clinical trial enrichment. Despite the promising predictive performance of recent longitudinal forecasting methods, they remain largely data-driven, struggle with irregularly sampled, incomplete longitudinal data and often neglect established disease biology, leading to biologically implausible trajectories. To address this, we propose a biologically constrained continuous-time framework for long-horizon cognition forecasting from limited baseline observations. The proposed method models the complete amy-loid–tau–vascular–neurodegeneration–cognition (ATVNC) cascade using hierarchical Neural ODEs with biologically motivated monotonicity constraints. Each pathological stream is governed by a dedicated Neural ODE initialized from irregular longitudinal observations using a GRU-D encoder, capturing intrinsic disease evolution while being modulated by directed upstream pathological influences. A bounded cognition readout ensures physiologically valid cognitive score (MoCA) predictions, while teacher–student knowledge distillation improves learning from sparse longitudinal supervision. Evaluated on the ADNI dataset, the proposed framework achieves a long-horizon extrapolation MAE of 2.06 on 188 held-out participants while eliminating biologically implausible trajectory violations. It further demonstrates robust zero-shot cross-cohort generalization on OASIS-3 (MAE 2.68 on 300 participants), with fine-tuning improving MAE to 1.90. The model also supports prognostic enrichment for Alzheimer’s clinical trials, achieving up to 2.70 × enrichment over the cohort base rate. These results demonstrate that embedding biological disease mechanisms within continuous-time deep learning improves the accuracy, biological plausibility, and clinical utility of long-horizon cognitive forecasting. The code is publicly available at: https://github.com/PonDeepika/BEACON.

## 1. Introduction

Alzheimer’s disease (AD) accounts for 60–80% of the more than 55 million dementia cases worldwide, a burden projected to triple by 2050 in the absence of disease-modifying therapies (Xiang et al., 2026). Its underlying pathology unfolds over decades as an ordered cascade of molecular and structural changes: extracellular amyloid-β accumulation, intraneuronal tau propagation along connectome pathways, cerebrovascular injury, progressive neurodegeneration, and ultimately cognitive decline (Hampel et al., 2021; Knopman et al., 2021). These processes are neither independent, rather, they interact through nonlinear dynamics that vary substantially across individuals and disease subtypes, rendering the reliable long-horizon forecasting of individual cognitive trajectories a fundamentally challenging problem.

Accurate long-horizon forecasting of cognitive decline carries direct clinical value. Reliable subject-specific predictions enable refined patient stratification for clinical trials, support individualised prognosis and timely therapeutic intervention, particularly at the pre-dementia mild cognitive impairment (MCI) stage where treatment effects are greatest (Aisen et al., 2017). To this end, large scale longitudinal cohort studies, principally the Alzheimer’s Disease Neuroimaging Initiative (ADNI) (Petersen et al., 2010) and the Open Access Series of Imaging Studies (OASIS-3) (LaMontagne et al., 2019), have generated richly annotated multimodal datasets encompassing structural Magnetic Resonance Imaging (MRI), amyloid and tau Positron Emission Tomography (PET), cerebrospinal fluid (CSF) biomarkers, diffusion imaging, and serial neuropsychological assessments. These resources provide an unprecedented substrate for data-driven disease progression modeling, yet exploiting them is non-trivial: participants exhibit heterogeneous biomarker availability, irregular and subject-specific visit intervals, and substantial longitudinal missingness. Standard supervised learning pipelines are ill-equipped to handle above non-trivial challenges.

A growing body of longitudinal forecasting work has sought to address these challenges using recurrent models with temporal decay (Che et al., 2018; Aghajanian et al., 2025), imputation-aware sequence models (Xu et al., 2022), and temporal attention-based encoders (Shukla and Marlin, 2021; Al Olaimat et al., 2024) that operate directly on irregular observed visit sequences. On the other hand, continuous-time formulations including Latent ODEs (Rubanova et al., 2019), Neural CDEs (Kidger et al., 2020), and hybrid ODE–recurrent models (Bossa and Sahli, 2023; Zanin et al., 2025) represent disease progression as a continuous latent dynamical process, rather than modeling only transitions between observed visits, and have demonstrated competitive accuracy on benchmarks such as TADPOLE (Marinescu et al., 2019). Despite this progress, a fundamental limitation persists across these approaches: they learn disease trajectories from statistical regularities in data alone, without incorporating the known directionality, temporal ordering, or inter-stream interaction structure of AD pathology. Consequently, learned trajectories can be biologically implausible, for instance, exhibiting declining amyloid or tau burden and increasing neuroanatomical volumes in the absence of any disease-modifying intervention. As a result, models generalize poorly to unseen forecast horizons or cohorts with different missingness profiles.

Conversely, a complementary body of work has encoded biological structure as its starting point. Event-based models (Fonteijn et al., 2012; Young et al., 2021), disease progression scoring methods (Jedynak et al., 2012; Donohue et al., 2014), and Bayesian mixed-effects trajectory models (Lorenzi et al., 2019) represent population-level disease timelines, while network diffusion (Raj et al., 2012) and connectome-based ODE systems (Wen et al., 2025) propagate molecular pathology spatially across the structural connectome. These approaches typically model only a narrow subset of the AD cascade (most focus on amyloid and tau in isolation), cannot readily ingest irregular multimodal longitudinal data, and are not designed for end-to-end forecasting of downstream cognitive outcomes. Existing work thus occupies two poles: flexible but biologically unconstrained data-driven models at one extreme, and interpretable but clinically limited mechanistic models at the other, without a framework to successfully unify both.

An equally significant but underappreciated limitation concerns the choice of cognitive endpoint. Majority of data-driven forecasting studies target the Mini-Mental State Examination (MMSE) (Zanin et al., 2025) or the Alzheimer’s Disease Assessment Scale-Cognitive Subscale (ADAS-Cog) (Marinescu et al., 2019; Ma et al., 2026). However, MMSE exhibits a well-documented ceiling effect that substantially limits sensitivity during early MCI (the stage where intervention is most beneficial) because scores cluster within a narrow normal range and change minimally even as clinically meaningful cognitive erosion occurs (Nasreddine et al., 2005). Although, ADAS-Cog avoids this floor insensitivity but is primarily a research instrument, administered in specialist settings, and is not routinely collected in clinical practice. In contrast, Montreal Cognitive Assessment (MoCA) offers superior sensitivity across the full MCI spectrum (Nasreddine et al., 2005), a wider effective dynamic range, and widespread global adoption in outpatient and memory clinic settings. Despite this clinical relevance, MoCA is often just treated as an outcome measure in observational cohorts and has received almost no attention as a prediction target for longitudinal machine learning models, representing both a gap in the literature and an opportunity for immediate translational impact.

Taken together, three interrelated gaps remain unaddressed: (i) no existing framework embeds established AD pathological structure as a structural prior within a flexible continuous-time model capable of handling irregular, incomplete multimodal data; (ii) biological constraints such as monotonicity, cascade ordering, and inter-stream coupling, necessary for clinically plausible long-horizon extrapolation, are absent from data-driven approaches; and (iii) the pervasive use of cognitively insensitive or clinically inaccessible targets has left MoCA-targeted long-horizon forecasting an open problem.

To address these gaps, we propose a biologically constrained continuous-time dual-encoder teacher–student framework, for long-horizon cognition (MoCA) forecasting. First grounded in the Jack et al. (Jack et al., 2013) biomarker cascade and the Amyloid (A)/Tau (T)/ Neurodegeneration (N) framework (Jack Jr et al., 2018), the proposed work extends the classical ATN axis into a dynamic predictive framework by introducing vascular injury (V), capturing both intrinsic cerebral small vessel disease (CSVD) and amyloid-, tau-exacerbated vascular pathology, and cognition (C) as the terminal clinical endpoint, forming the proposed ATVNC cascade. To computationally model this cascade, we introduce BEACON (Biologically-constrained Evolutionary ATVNC framework for long-horizon COgnitioN forecasting), which represents continuous-time disease progression through five hierarchically coupled, monotonicity-constrained latent streams evolved by a structured latent Neural ODE (NODE) (Rubanova et al., 2019) whose vector field explicitly encodes directed inter-stream interactions. Irregular and incomplete longitudinal observations within a 24-month clinical window are encoded by a GRU-D encoder (Che et al., 2018), with biologically conditioned stream-specific routing networks. We incorporate age, APOE genotype, DTI measures, and microbleed burden as stream-relevant context, initializing latent states under per-stream auxiliary supervision. Deep microbleeds are treated as markers of hypertensive CSVD, and lobar microbleeds for cerebral amyloid angiopathy (CAA)-related pathology (Liu et al., 2025e) associated with the amyloid axis. To bridge the limited clinical observation window and richer training histories, a teacher encoder observing complete follow-up supervises both the latent initialization and predicted trajectories of the deployment-restricted student. We evaluate BEACON through state-of-the-art comparison, horizon-stratified {0 →12, 12 →24, 24 → 36, 36 → 48, 48 →60 and > 60 months}, clinical subgroup analysis, robustness to observation window length, biological plausibility validation, and systematic ablation of streams, loss components, and architectural choices.

The primary contributions of this work are as follows:

- **Biologically constrained ATVNC latent ODE**. We design a biologically constrained latent NODE whose vector field models five interacting biological streams through a learnable directed coupling hierarchy reflecting the ATVNC disease cascade. To our knowledge, this is the first end-to-end continuous-time framework to explicitly embed the complete ATVNC hierarchy within latent dynamics for multimodal long-horizon cognitive forecasting.
- **Direction-aware monotonicity constraints**. We enforce stream specific monotonicity, guaranteeing that accumulating (A, T, V) and atrophic (N) processes evolve in their prescribed biological directions at all forecast horizons, eliminating a class of trajectory violations that result in unconstrained data-driven baselines.
- **Stream-aware latent partitioning with auxiliary supervision**. We introduce stream-specific routers initializing biologically disjoint latent streams conditioned on static biomarkers with per-stream auxiliary regression heads preserving biomarker-aligned representations throughout the ODE evolution.
- **Teacher-student dual encoder for long-horizon forecasting**. Our knowledge distillation framework consists of a teacher encoder that observes complete longitudinal histories and supervises both the latent initialization and predicted trajectories of a student encoder, enabling robust long-horizon MoCA forecasting from limited (24-month) observation windows.
- **MoCA as a clinically grounded forecasting target**. We present the first deep learning framework to our knowledge for long-horizon MoCA forecasting, demonstrating superior early-stage sensitivity over MMSE-targeted baselines.
- **Systematic experimental validation;** We establish the state-of-the-art MoCA forecasting performance, demonstrate the biological plausibility of the learned trajectories, and quantify the marginal contribution of each ATVNC stream and methodological components to predictive accuracy.

## 2. Related Work

### 2.1. Longitudinal learning under irregular sampling and missingness

Clinical longitudinal data are characterized by irregular visit intervals and pervasive missingness that invalidate standard fixed-step recurrent architectures. Hence, handling irregular and missing observations remains a central challenge in clinical longitudinal modeling. Discrete-time recurrent methods handle irregular sampling by incorporating temporal information through modified recurrent units. For instance, Time-aware LSTM (T-LSTM) (Baytas et al., 2017) models decay memory states across irregular visit intervals, Gated Recurrent Units-Decay (GRU-D) jointly models irregular sampling and missingness using exponential decay and masking (Che et al., 2018). More recently, attention- and transformer-based models, such as Multi-Time Attention Networks (mTAN) (Shukla and Marlin, 2021) and Timely Generative Pre-trained Transformer (Time-lyGPT) (Song et al., 2024), have demonstrated strong performance on irregular clinical time-series through temporal attention and learned representations.

However, these approaches remain discrete-time sequence models and do not explicitly model continuous disease evolution for forecasting at arbitrary time points. In contrast, continuous-time latent variable models, including Neural ODEs (Chen et al., 2018; Dupont et al., 2019), ODE-RNNs (Rubanova et al., 2019), GRU-ODE-Bayes (De Brouwer et al., 2019), Neural CDEs (Kidger et al., 2020), Neural RDEs (Morrill et al., 2021) and continuous-time Transformer architectures such as ContiFormer (Chen et al., 2023b), parameterize temporal evolution as neural differential systems, naturally accommodate irregular sampling without interpolation. Yet, none of these encoding strategies impose biological structure on the learned representations.

### 2.2. Data-driven AD progression forecasting

Modeling of AD progression has evolved from probabilistic staging toward continuous-time trajectory learning. Data-driven forecasting of AD progression has been systematically benchmarked through the TAD-POLE challenge (Marinescu et al., 2019), which established a broad comparison of statistical and machine learning methods. Event-based models represent progression as an ordered sequence of biomarker abnormality events (Fonteijn et al., 2012), later extended by Subtype and Stage Inference (SuStaIn) to jointly recover progression subtypes and stages (Young et al., 2021). In parallel, disease progression scores methods (Jedynak et al., 2012; Donohue et al., 2014) locate each patient on a single shared biomarker timeline, while Bayesian mixed-effects models (Lorenzi et al., 2019) infer individualized trajectories with uncertainty estimates. Disease course mapping methods (Koval et al., 2021) warp a shared trajectory template through per-subject time reparameterization. These methods assume smooth population-level progression and do not readily accommodate irregularly observed multimodal inputs or produce direct cognitive outcome forecasts.

Deep longitudinal forecasting models learn temporal disease dynamics directly from sequential observations. Recurrent architectures have been applied to forecast biomarker and cognitive trajectories (Nguyen et al., 2020), with extensions handling irregular sampling and missingness (Aghajanian et al., 2025; Al Olaimat et al., 2024) and learnable multimodal imputation (Xu et al., 2022). Continuous-time disease progression has been modeled using Bayesian ODE formulations (Bossa and Sahli, 2023) and Neural ODE-based approaches with recurrent (Zanin et al., 2025) and convolutional (Sharma et al., 2025) encoders, alongside latent flow models for longitudinal MRI progression such as ImageFlowNet (Liu et al., 2025b). At the other extreme, purely cross-sectional methods predict clinical outcomes from a single baseline examination (Ma et al., 2026; Basaia et al., 2019) but discard longitudinal disease evolution and cannot model individualised progression trajectories. Across both paradigms, methods are primarily optimised for statistical accuracy without incorporating established disease biology, limiting biological interpretability and often yielding physiologically implausible trajectory predictions. Furthermore, existing longitudinal approaches target diverse endpoints - MMSE (Bossa and Sahli, 2023; Sharma et al., 2025; Zanin et al., 2025), ADAS-Cog (Aghajanian et al., 2025), AD conversion (Nguyen et al., 2020), and MRI progression (Liu et al., 2025b), while longitudinal MoCA forecasting remains largely unexplored.

### 2.3. Biologically constrained models of AD Pathology

Another line of work incorporates established disease biology as an explicit structure. The hypothetical biomarker cascade of Jack et al. (Jack et al., 2013) describes the characteristic temporal ordering of AD pathophysiology, from amyloid accumulation preceding tau propagation, neurodegeneration, and cognitive decline, providing the biological rationale for the A/T/(N) classification framework (Jack Jr et al., 2018) that stratifies individuals by biomarker abnormality status. These established dependencies motivate the structural priors encoded in our ATVNC cascade. Network diffusion models (Raj et al., 2012) represent the brain as a structural connectome and predict spatio-temporal spreading of misfolded pathology along white-matter fibre connections. More recent connectome-based ODE models (Wen et al., 2025) fit individualized amyloid and tau trajectories aligned onto a common disease timeline. Additionally, cerebrovascular disease and white matter hyperintensity (WMH) burden represent an important and partially independent contributor to cognitive decline in AD (Lo et al., 2012). Evidence for amyloid-angiopathy-driven WMH accumulation (Chen et al., 2006) supports the A → V coupling in our model, while the V →N pathway reflects emerging evidence of CSVD-related neurodegeneration (Wardlaw et al., 2015).

The above mechanistic models, despite being physiologically interpretable, are restricted to a small set of molecular markers, rely on a predefined coupling graph, and cannot ingest irregular multimodal longitudinal data or forecast downstream cognitive outcomes end-to-end. Existing work occupies two extremes: flexible but biologically unconstrained data-driven dynamics, or interpretable but inflexible mechanistic structure. No framework successfully unifies both within a trainable model capable of end-to-end cognitive trajectory forecasting from heterogeneous clinical data.

### 2.4. Knowledge distillation and teacher-student frameworks in longitudinal forecasting

Knowledge distillation (KD) (Hinton et al., 2015) transfers information from an information-rich teacher to a constrained student model, and has emerged as an effective paradigm for improving long-horizon time series forecasting under sparse observations and missing data (Ni et al., 2026). Beyond model compression, recent approaches exploit teachers with privileged temporal context or richer observation windows to transfer informative representations to student models (Liu et al., 2022). Representative methods include KD through time (Gunasekaran et al., 2024), which distils knowledge across forecasting tasks; Di-Long (Das et al., 2024), which transfers knowledge from short-to long-horizon trajectory prediction; DE-TSMCL (Gao et al., 2024), which combines distillation with contrastive temporal representation learning; TimeKD (Liu et al., 2025a), which exploits privileged information for multivariate forecasting; and TimeDistill (Ni et al., 2026), which demonstrates that large temporal models can be distilled into lightweight networks while preserving long-horizon performance. In the AD domain, KD has been applied primarily to missing-modality learning from incomplete imaging inputs (Chen et al., 2023a; Liu et al., 2025c,d), leaving the use of teacher-student learning from sparse, irregular longitudinal clinical observations largely unexplored for continuous cognitive trajectory prediction.

### 2.5. Cognitive endpoint/target selection in AD studies

Most AD modeling studies, including the TAD-POLE challenge, have predominantly adopted ADAS-Cog (Bossa and Sahli, 2023; Marinescu et al., 2019; Ma et al., 2026), Clinical Dementia Rating Sum of Boxes (CDR-SB) or global ordinal CDR transitions (Young et al., 2021) largely due to its historical role as a standard cognitive outcome and dense temporal availability in ADNI. Also, many recent data-driven models continue to use MMSE for its ubiquitous tracking across clinical centers (Zanin et al., 2025). However, conventional reliance on MMSE have disadvantage of aggressive *ceiling effect* (Franco-Marina et al., 2010; Luzzi and Snowden, 2026) that obscures cognitive breakdown during early MCI stages, the window when intervention is the most impactful. Although ADAS-Cog is more sensitive than MMSE, it remains primarily a clinical trial outcome measure requiring standardized administration by trained examiners, limiting its practicality for routine clinical use (Cecato et al., 2016). In contrast, MoCA has been underutilized despite extensive clinical evidence that it provides greater sensitivity to early MCI (Pinto et al., 2019). As it is widely utilized in routine clinical practice and tracked within longitudinal cohorts like ADNI and OASIS, MoCA has the potential to serve as a highly relevant and appropriate endpoint for personalized longitudinal progression models, despite its relative data sparsity.

## 3. Materials and methods

### 3.1. Dataset details

This study uses multimodal longitudinal data from ADNI phases 1, GO, 2, 3, and 4 studies (Jack Jr et al., 2008; Petersen et al., 2010) and OASIS-3 study (LaMontagne et al., 2019), two large publicly available cohorts spanning CN, MCI, and AD individuals across multiple clinical visits with multimodal assessments acquired over several years.

#### ADNI

Data used in the preparation of this article were obtained from the Alzheimer’s Disease Neuroimaging Initiative (ADNI) database (adni.loni. usc.edu). The ADNI was launched in 2003 as a public-private partnership with a goal to test whether serial MRI, PET, other biological markers, and clinical and neuropsychological assessment can be combined to measure the progression of MCI and early AD. We utilize pre-computed imaging and fluid biomarkers from centralized ADNI processing laboratories, ensuring standardized harmonization across acquisition sites. Amyloid burden is quantified on the Centiloid scale via cortical standardized uptake value ratios (SUVR) transformation by the UC Berkeley PET Core. Similarly, CSF phosphorylated tau-181 (pTau181) is measured using the Roche Elecsys immunoassay by the University of Pennsylvania Biomarker Core. Additional biomarkers comprise whole-brain morphometric measures derived from T1-weighted MRI using FreeSurfer (version 4.3 for ADNI-1 1.5T data; 5.1 for ADNI-GO/2 and ADNI-1 3T data; 6.0 for ADNI-3 3T data; and 7.4 for ADNI-4), total WMH volume, white matter diffusion tensor imaging (DTI) metrics comprising fractional anisotropy (FA), mean diffusivity (MD), axial diffusivity (AD), and radial diffusivity (RD), and cerebral microbleed (CMB) counts in deep and lobar regions.

#### OASIS

We use the OASIS-3 (https://www.nitrc.org/projects/oasis3/) cohort as an independent external validation dataset. Available biomarkers in OASIS-3 include Centiloid-scale amyloid burden, FreeSurfer-derived morphometrics (versions 5.0–5.3), and MoCA scores. WMH burden is estimated from FLAIR MRI using FSL TrUE-Net (Sundaresan et al., 2021b,a, 2022), which has demonstrated high reliability and low false-positive rates in independent validation (Strain et al., 2024). CSF pTau181, DTI metrics, and CMB measures are unavailable in this cohort and are treated as missing throughout.

#### Inclusion and partitioning

Inclusion requires at least one measurement of MoCA, amyloid, and CSF pTau181 (ADNI) or MoCA and amyloid (OASIS), with a minimum of two measurements for at least one primary biomarker, even if acquired asynchronously. This yields final cohorts of 1,235 ADNI and 649 OASIS participants. Partitioning is performed strictly at the participant level for both ADNI and OASIS to prevent data leakage; participants with three or fewer total MoCA visits are assigned exclusively to the training set. As expected for any longitudinal observational studies, both cohorts exhibit irregular follow-up intervals and substantial biomarker missingness; the average number of visits and biomarker-specific missingness patterns are shown in Figure 1. The final ADNI cohort comprises an average of 8.70 ± 5.23 visits per participant over a mean follow-up of 73 months, whereas the OASIS-3 cohort comprises 10.64 ± 5.71 visits per participant over a mean follow-up of 109 months.

**Figure 1:**
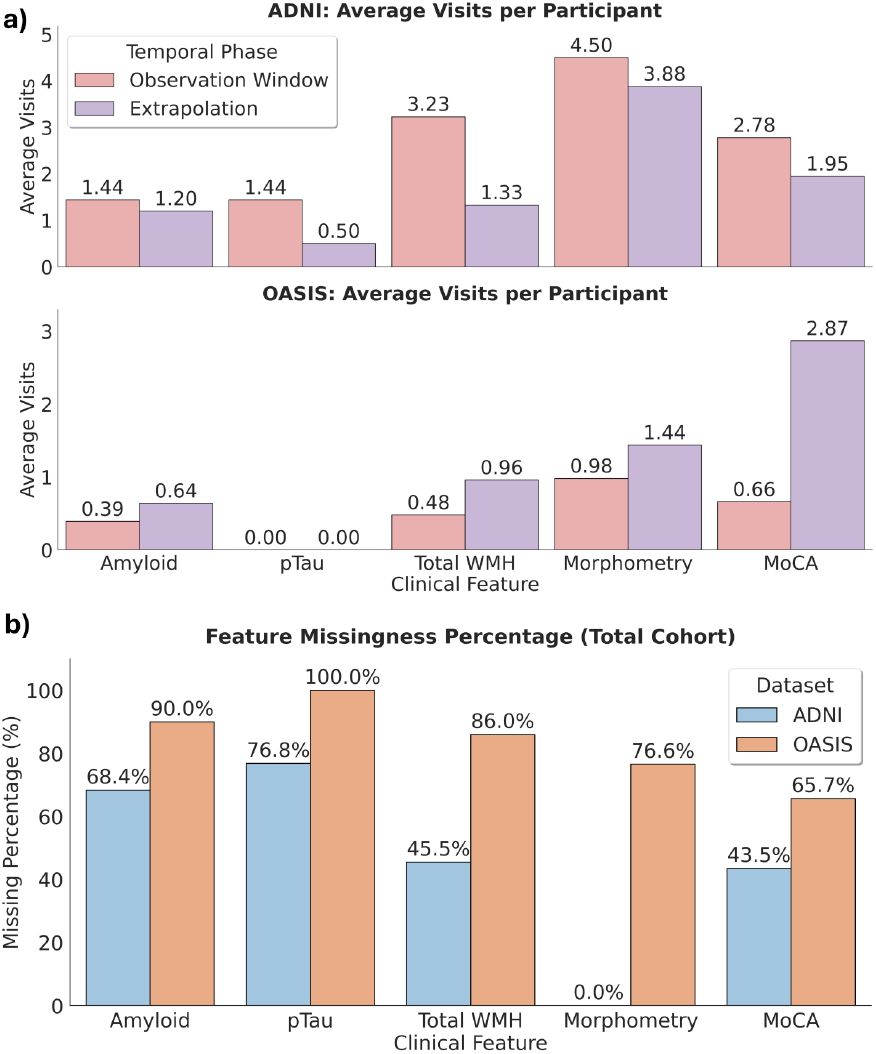
Longitudinal data availability and biomarker completeness in the ADNI and OASIS cohorts. (a) Average number of visits per participant across the observation window, extrapolation window, and the complete longitudinal follow-up for each cohort. (b) Percentage of missing observations for each longitudinal biomarker, computed with respect to the total number of visits in each cohort.

### 3.2. Surrogate biomarkers of ATVNC streams

Each ATVNC axis is instantiated using clinically established biomarkers available across both cohorts. The *A* stream is represented by amyloid PET burden, quantified as cortical SUVR referenced to the whole cerebellum and standardized to the Centiloid scale via the UC Berkeley PET Core pipeline (Section 3.1), giving a harmonized, radiotracer-independent measure of amyloid pathology. The *T* stream is represented by CSF pTau181, an established fluid biomarker of AD-related tau pathophysiology. The *V* stream is represented by total WMH volume (provided by the UC Davis Imaging Core for ADNI; estimated via TrUE-Net (Sundaresan et al., 2021b) for OASIS as described in Section 3.1), a robust marker of CSVD associated with cognitive decline through disruption of structural white matter connectivity (Hu et al., 2021). The *N* stream is represented by structural morphometric measures derived from T1-weighted MRI via FreeSurfer processing. To obtain a compact, predictively relevant representation, stable morphometric features are identified through patient-level bootstrap LASSO regression (200 resamples) on the ADNI training and validation set, using intracranial volume (ICV) normalized regional brain volumes at baseline to predict each participant’s first available follow-up MoCA score beyond 24 months. Features are ranked by selection frequency across bootstrap resamples as a stability score. Middle temporal gyrus and hippocampal volumes emerge as the most stable predictors (stability score = 1.0), consistent with AD’s characteristic medial temporal atrophy (Convit et al., 2000; Visser et al., 2002). Ventricular enlargement and CSF expansion, the next highest-ranked features, capture the same ex-vacuo atrophic process and are merged into a single CSF compartment. Lower-ranked structures are excluded as they provide redundant information without improving predictive performance. Consequently, *N* stream is decomposed into three sub-streams: hippocampal atrophy (*H*), middle temporal gyrus atrophy (*M*), and CSF expansion (*F*). The *C* stream is assessed by MoCA, selected for its sensitivity across the MCI spectrum and widespread clinical availability (Nasreddine et al., 2005).

### 3.3. Problem Formulation

Let 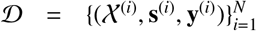 denote a longitudinal cohort of *N* participants. For subject *i*, the multimodal longitudinal sequence observed within the 24-month clinical observation window is 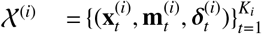 where *K*_*i*_ is the number of visits, **x**_*t*_= [*A*_*t, t*_ *T*_*t*_ *V*_*t*_, *M*_*t*_, *H*_*t*_, *F*_*t*_, *C*_*t*_] ∈ R^*D*^ is the multimodal biomarker observation vector at visit *t*, with *A, T*, *V, M, H, F* and *C* denoting amyloid burden, CSF pTau181, WMH volume, middle temporal gyrus volume, hippocampal volume, CSF compartment volume, and MoCA score, respectively; **m**_*t*_ ∈ {0, 1}^*D*^ denotes the missingness mask, and δ_*t*_ ∈ R^*D*^ represents the elapsed time since the last valid observation of each feature, accommodating irregular and asynchronous sampling across modalities.

Each subject is associated with a static covariate vector **s** = [Age, APOE, DTI, CMB, ICV, τ], comprising time-invariant variables or biomarkers too sparsely observed for longitudinal modeling, where ICV denotes intracranial volume and τ encodes observation-month tokens indicating when each static biomarker was last measured (τ =− 3 in z-score space when absent). Biologically relevant stream-specific context vectors are selected as **s**_*X*_ ⊆ **s**, *X* ∈{*A, T, V, H, M, F, C*}, conditioning each biological stream on only its biologically relevant static covariates, as detailed in Section 3.6.

The future observations beyond the observation cutoff *T*_obs_ are denoted as 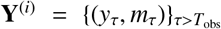, where *y*_τ_ includes the primary outcome (MoCA) and auxiliary biomarker targets at irregularly spaced future times τ.

#### Objective

Given the observations within the clinical window O = {X, **s**}, the goal is to learn a mapping

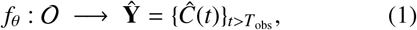

that produces continuous-time MoCA trajectory predictions *Ĉ*(*t*) ∈ [0, 30] for *t* > *T*_obs_, subject to two structural constraints: (i) direction-aware monotonicity on accumulating and atrophic biological streams, and (ii) clinical range enforcement on MoCA predictions throughout the forecast horizon.

#### Teacher-student split

During training, a teacher encoder additionally observes the complete longitudinal sequence **Y**_full_ = *Χ* ∪ **Y** spanning all available follow-up visits (mean: 46 ± 34 months, range: [6, 180] months across the training cohort). At inference, only the student encoder is active, observing *Χ* alone within the 24-month window.

### 3.4. Framework Overview

The proposed framework comprises four sequentially coupled components, illustrated in Figure 2. Given the longitudinal observations and static covariates within the clinical observation window O = {*Χ*,**s** } : *(i)* a missingness-aware temporal encoder summarizes the irregularly sampled longitudinal biomarker history (*Χ*) into a compact patient-specific global representation; *(ii)* a stream-specific router projects this representation, augmented with biologically relevant static context, into seven biologically partitioned latent initial states; *(iii)* a structured Neural ODE governs the continuous-time joint evolution of these states under two complementary biological priors; and *(iv)* stream-specific readout heads decode the resulting latent trajectories into future biomarker predictions and the primary cognitive outcome.

**Figure 2:**
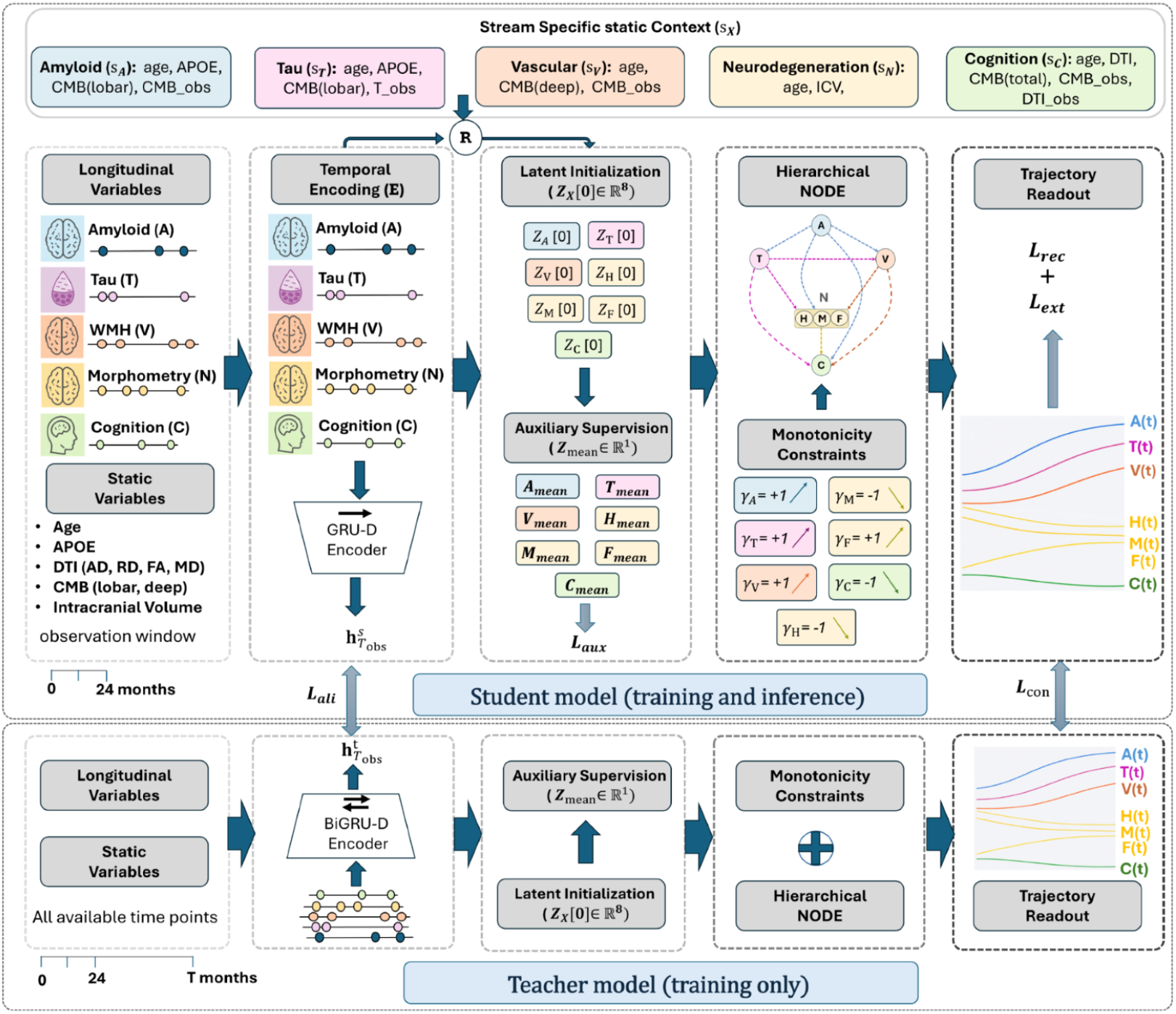
Overview of the proposed teacher–student BEACON architecture. A missingness-aware GRU-D encoder (E) generates a global patient representation (*h*_*T*obs_) from the observation window, which is routed(R) together with stream-specific static context (*s*_*A*_) to initialize stream-specific latent representations (*Z*_*x*_[0]). Auxiliary supervision (ℒ_aux_) is applied to each initialized latent state to encourage biomarker-specific representations. The latent streams are evolved by hierarchical Neural ODEs under biological progression constraints to predict longitudinal biomarker and cognitive trajectories, optimized using reconstruction (ℒ_*rec*_) and extrapolation (ℒ_*ext*_) losses. During training, a privileged BiGRU-D teacher encoder accesses the full longitudinal history, distilling knowledge to the student (with access to 24-month window) through latent alignment (ℒ_*ali*_) and consistency (ℒ_*con*_) losses. At inference, only the student branch is deployed.

Formally, the temporal encoder maps the observation sequence to a global latent representation, 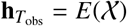. For each stream *X*, a stream-specific context vector is obtained as **ctx**_*X*_ = *g*(**s**_*X*_), where *g*(·) denotes the context projection network. The global latent representation is then combined with the corresponding stream-specific context to initialize the latent state of each stream,

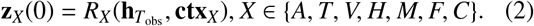

These states evolve jointly under the biologically constrained ODE,

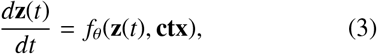

whose dynamics encode two structural priors. First, a learnable hierarchical coupling cascade following the directed ATVNC hierarchy allows upstream pathological processes to influence downstream mechanisms, directly instantiating the (Jack et al., 2013) biomarker cascade within the latent dynamics rather than as a post-hoc constraint. Second, direction-aware monotonicity constraints enforce biologically prescribed accumulation for amyloid, tau, WMH, and ventricular CSF expansion, and progressive decline for hippocampal and middle temporal gyrus volumes, guaranteeing clinically plausible trajectory extrapolation at arbitrary future horizons. Stream-specific readout heads *D*_*X*_ decode the evolved latent trajectories into biomarker predictions, providing auxiliary supervision signals, with the cognition stream producing the primary forecast,

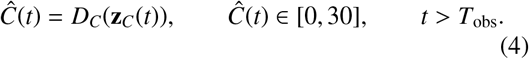

To bridge the 24-month clinical observation window available at deployment and the richer longitudinal histories available during training, the framework adopts a dual-encoder teacher-student strategy. A teacher encoder, observing the complete longitudinal follow-up sequence, supervises both the latent initialization and predicted trajectories of a student encoder restricted to the observation window. At inference, the teacher is completely discarded and only the student is deployed.

### 3.5. Temporal Encoding of Longitudinal Biomarkers

The temporal encoder *E*(·) maps each patient’s longitudinal biomarker history within the *T*_obs_ = 24-month window into a compact global representation 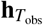, requiring robustness to missing values and irregular sampling intervals.

#### Expanded input representation

Although the downstream ODE operates on the 7-dimensional biomarker vector **x**_*t*_ (Section 3.3), the encoder ingests an expanded vector 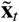 in which the three neurodegeneration biomarkers (*M*_*t*_, *H*_*t*_, *F*_*t*_) are replaced by the top 10 stable morphometric volumes identified via bootstrap LASSO regression (described in Section 3.2). This allows the encoder to exploit more comprehensive anatomical context from structural MRI.

#### GRU-D encoder

GRU-D (Che et al., 2018) is adopted over ODE-based encoders (Rubanova et al., 2019; Kidger et al., 2020) for its established performance on clinical sequences with high missingness, computational efficiency, and feature-level decay suited to the modality-specific missingness structure of ADNI and OASIS. The decay rate for feature *d* at visit *t* is γ_*t*,*d*_ = exp(−max(0, **W**_γ_ δ_*t*,*d*_ + *b*_γ_)), with missing values imputed as a decay-weighted interpolation between the previous hidden state and the training-set empirical mean 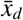. The hidden state is updated as:

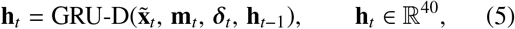

where the GRU-D encoder contains 8,388 trainable parameters. The hidden dimension is selected by validation-set search over {16, 32, 40, 64, 128}, balancing reconstruction against overfitting given the modest cohort size and the low intrinsic dimensionality of the biomarker set. The final hidden state 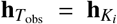 yields the global, subject-specific compact latent representation summarizing the clinical history, serving as the shared initialization input to all streams via routing networks.

The teacher encoder shares this architecture but maintains independent parameters and observes the complete longitudinal sequence, as detailed in Section 3.9.

### 3.6. Stream-Specific Latent Initialisation

The global representation 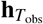 produced by the GRUD encoder is augmented with the stream-specific static context vector (**ctx**_*X*_), derived from the static covariates (s). The representation 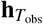 is projected into biologically partitioned latent initial states via stream-specific routing networks. Each stream’s initial state is obtained as

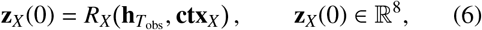

where *R*_*X*_(·) is a two-layer MLP: a linear layer followed by ReLU and dropout, then a linear output layer with no activation, leaving **z**_*X*_(0) unconstrained in the latent space. The stream-specific context vector **ctx**_*X*_ is obtained by projecting stream-relevant static features **s**_*X*_ through an affine layer with ReLU activation,

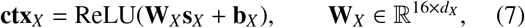

where the stream-specific feature subsets **s**_*X*_ ∈ ℝ^*dX*^ are:

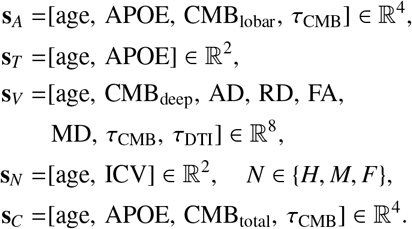

The context assignments are biologically grounded: APOE4 and lobar CMBs condition the amyloid stream; DTI scalars and deep CMBs condition the vascular stream to capture white-matter integrity and CSVD burden; ICV scales the *N* stream to account for head size variation in volumetric measures.

#### Auxiliary supervision

To regularize the latent initialization and anchor each stream to its underlying biological burden, a lightweight auxiliary linear head is attached to each initial latent state **z**_*X*_(0) to predict the observed stream mean µ 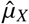 within the observation window,

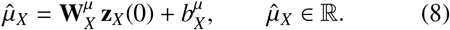

Unlike the forecasting losses, which provide gradients only through ODE integration, the auxiliary head provides a direct supervision signal at t=0, thereby stabilizing latent initialization and encouraging an initial condition that is informative for subsequent long-horizon forecasting. Consequently, the auxiliary prediction task guides the initial latent state towards representing the subject’s underlying biological burden, while the primary forecasting objective remains the dominant learning signal.

Thus, the latent-initialization module contains 7,784 trainable parameters, comprising 448 in the static-context projections and 7,336 in the stream-routing network (seven per-stream MLPs). Auxiliary stream-mean heads add only 63 additional parameters and are used exclusively during training, for a total of 7,847 parameters during training.

### 3.7. Biologically constrained ODE dynamics

The ODE vector field *f* (*t*, **z**) implements the ATVNC cascade with a hierarchically structured formulation shared across all channels, differing only in their parent sets and biological sign. Every channel evolves under a common structured vector field:

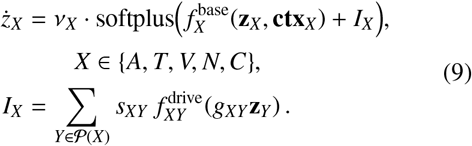

where 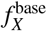 is a two-layer tanh-bounded MLP with hidden dimension 16, capturing self-dynamics conditioned on static context **ctx**_*X*_; 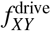 is a single linear layer with tanh activation encoding the directed influence of parent stream *Y* on *X*; *g*_*XY*_ is a learnable scalar pathway gate; and *s*_*XY*_ > 0 is a learnable amplitude scale parameterized via softplus. The biological sign *ν*_*X*_ +1, 1 enforces accumulation (*ν*_*A*_ = *ν*_*T*_ = *ν*_*V*_ = *ν*_*F*_ = +1) or monotone decline (*ν*_*H*_ = *ν*_*m*_ = *ν*_*C*_ = − 1). The softplus guarantees a non-negative magnitude, ensuring each stream is strictly monotone in its biologically prescribed direction.

#### Directed pathway structure

The parent sets (*X*) encode the ATVNC coupling hierarchy:

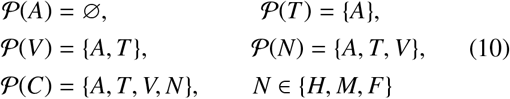

Longitudinal amyloid PET studies demonstrate monotonic accumulation with an early acceleration phase that saturates at higher burden (Villemagne et al., 2013; Jack Jr et al., 2013); rather than imposing a fixed logistic form, the tanh-bounded base network learns this profile directly from data, while the softplus and *ν*_*A*_=+1 guarantee monotone accumulation. Tau accumulation is modeled as unidirectionally driven by amyloid, consistent with the amyloid cascade hypothesis (Hardy and Higgins, 1992) and supported by longitudinal ADNI analyses in which *A*β → τ coupling significantly out-performs the reverse direction (Raj et al., 2025). A self-dynamics term additionally captures autonomous tau propagation independent of concurrent amyloid levels. The vascular stream receives directed input from both amyloid and tau: the *A* → *V* pathway reflects amyloid angiopathy-driven cerebrovascular impairment (Vernooij et al., 2008), while the *T* → *V* pathway is motivated by recent longitudinal evidence of a directional hierarchy between AD pathology and WMH progression (Alban et al., 2023; McAleese et al., 2015; Kamal and Dadar, 2026). We acknowledge that the precise directionality of *A*/*T* -WMH coupling remains debated, hence both *A* and *T* pathways are therefore implemented as soft priors modulated by learnable gatesv *g*_*XY*_ (Eqn. 9), which the model may suppress if unsupported by data. By embedding this directional hierarchy into the model, we allow the data to flexibly drive the strength of these downstream pathological pathways. *N*, and *C* follow as progressively downstream channels as per the parent sets above. Accordingly, the NODE module contains 15,341 trainable parameters distributed across the seven stream-specific dynamics modules as A:1,064, T:1,618, V:1,875, H/M/F:2,132 each, and C:4,388, with parameter counts increasing with the number of upstream inputs.

### 3.8. ODE integration and trajectory readout

#### Integration

Given the partitioned initial state **z**(0) produced by the routing networks, the vector field in Eq. (9) is integrated forward using the adaptive Dormand-Prince (`dopri5`) solver implemented in `torchdiffeq`, with relative and absolute error tolerances of 1 × 10^−7^ and 1 × 10^−9^, respectively. Because **z**(*t*) is continuous, it is evaluated directly at each patient’s irregularly spaced visit times without interpolation, naturally accommodating non-uniform longitudinal sampling.

#### Biomarker readout heads

The evolved latent trajectories **z**_*X*_(*t*), *X* * *C*, are decoded into biomarker predictions via stream-specific monotone readout heads, each implemented as a two-layer MLP with hidden dimension 16, softplus-parameterized non-negative weights, and tanh activations. Non-negative weights ensure the decoded biomarker is monotonically non-decreasing with respect to its latent state. The direction of monotonicity is inherited from *ν*_*X*_, so amyloid, pTau, WMH, and CSF expansion increase over time while hippocampal and middle temporal gyrus volumes decrease. The tanh activation induces smooth saturation, preventing unbounded growth at long horizons without requiring manually specified output bounds.

#### Bounded MoCA readout

MoCA score, a measure of cognition occupies a fixed clinical range within [0, 30]. The cognition latent **z**_*C*_ is decoded by a distinct bounded head that enforces the admissible MoCA range [0, 30] by construction. The head applies an affine projection of **z**_*C*_ and scaled logistic function to produce a MoCA prediction in original clinical units,

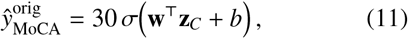

where **w**, *b* are learnable and σ(·) is the logistic function.

The above equation guarantees 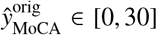 at arbitrary horizons. Unlike the biomarker heads, the sign of **w** is left unconstrained: while *ν*_*C*_= − 1 provides a physiologically grounded tendency toward decline, observed MoCA scores fluctuate and may transiently stabilize or improve due to therapy effects, measurement variability, and cognitive reserve.

Because regression losses are computed in *z*-score space, the bounded prediction is standardized prior to the loss as

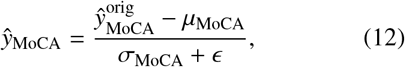

where (µ_MoCA_, σ_MoCA_) are computed on the training set only. At inference, Eq. (12) is inverted, recovering predictions on the native clinical scale. The readout module comprises 8,195 trainable parameters with 7,872 across the six monotone biomarker heads (1,312 each) and 323 in the bounded MoCA readout.

### 3.9. Student–Teacher Encoder Design

To enable robust long-horizon forecasting from limited early observations, we adopt a dual-encoder teacher–student framework.

#### Student encoder

The student encoder observes longitudinal biomarker measurements within the 24-month clinical observation window, producing the latent representation 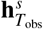. This is routed through the stream-specific networks to initialize the latent NODE states **z**^*s*^(0), which are evolved and decoded to yield the predicted trajectories 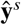.

#### Teacher encoder

The teacher employs a BiGRU-D encoder to exploit the both past and future observations within the full training sequence, producing a temporally richer supervisory signal. Given the complete longitudinal sequence available during training (mean: 46 ± 34 months), the teacher produces the information-rich representation 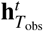, the initialization **z**^*t*^ (0), and trajectory predictions 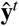. Access to future observations (that are unavailable to the student) enables the teacher to capture richer disease dynamics, making it a privileged supervisory signal.

#### Dense trajectory distillation

Rather than supervising the student only at sparse, irregularly sampled clinical visits, the teacher trajectory is evaluated at uniform one-month intervals and used as the distillation target throughout the full forecasting horizon. This dense temporal supervision provides smoother, less noisy guidance and encourages the student to learn rich disease dynamics from the 24-month observation window alone. The corresponding distillation losses are detailed in Section 3.10.

At inference, the teacher is discarded entirely and only the student encoder is deployed, reflecting the realistic clinical setting in which predictions must be made from the patient’s early observational history alone.

### 3.10. Composite Loss Function

The composite training objective is built from five terms targeting distinct aspects of the longitudinal learning problem, split into reconstruction, auxiliary initialization, and distillation objectives. All terms are computed over observed values using validity masks where appropriate.

#### Extrapolation and reconstruction losses

ℒ_ext_ denotes the mean squared error (MSE) between the predicted and observed biomarker trajectories over the extrapolation window. ℒ _rec_ denotes the MSE between observed biomarkers within the observation window and their reconstructions, obtained by evolving 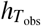 through a Neural ODE and decoding the resulting trajectory. To ensure that the optimization heavily prioritizes the primary clinical endpoint, we employ an explicit task-weighting.

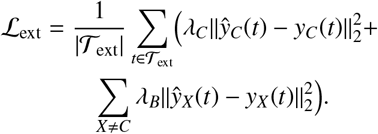

where *y*_*C*_ denotes primary MoCA stream (*C*), and {*X A, T, V, H, M, F, C*}. The reconstruction loss, ℒ_rec_ is formulated analogously over the baseline observation horizon _obs_ using the identical per-stream weight distribution.

#### Auxiliary initialization loss

ℒ_aux_ supervises each stream’s latent initial state by penalizing the MSE between the decoded stream mean prediction 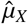 and the observed masked stream mean *µ*_*X*_ within the baseline window, employing uniform per-stream weights:

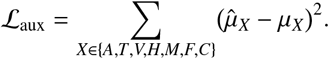

#### Distillation losses

The knowledge distillation framework encapsulates two complementary objectives grouped asℒ_distill_ = ℒ_con_ + ℒ_ali_. First, the latent alignment lossℒ_ali_ aligns the student latent representation to the information-richer teacher representation via an *ℓ*_1_ distance:

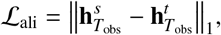

which encourages the restricted 24-month student window to recover the teacher’s full longitudinal encoding context.

Second, the trajectory consistency loss ℒ_con_ minimizes the MSE between student and frozen teacher trajectory predictions evaluated at uniform one-month intervals over the full extrapolation horizon [*T*_obs_, *T*_max_]:

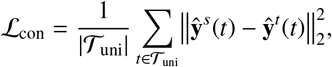

where *T*_uni_ denotes the set of uniformly spaced monthly evaluation times. Dense supervision across the full horizon provides smoother distillation targets and regularizes long-horizon forecasting against sparse data drift.

#### Composite objective

The student network is trained end-to-end by minimizing the weighted sum

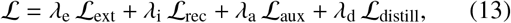

with λ_e_ = 1 (with λ_*C*_ = 3, λ_*B*_ = 0.5), λ_i_ = 1, λ_a_ = 2, and λ_d_ = 1 (withℒ_con_ = 0.5, ℒ_ali_ = 0.5), selected by grid search over {0.5, 1, 2, 3} on the validation fold. The elevated weight on MoCA reflects the primacy of MoCA as the clinical endpoint.

#### Training procedure

The teacher network is first trained independently using ℒ_ext_, ℒ_rec_, and ℒ_aux_ only, without the distillation terms. The teacher is subsequently frozen, and the student is trained on the full composite objective in Eq. (13).

### 3.11. Implementation Details

#### Preprocessing

Continuous features are *z*-score normalized using training-split statistics, with missing entries excluded from normalization statistics and indicated by binary observation masks. To reduce subject-specific baseline variability, decoded biomarker trajectories are anchored by applying a constant offset such that their mean over the observation window matches the corresponding observed mean, allowing the reconstruction loss to focus on longitudinal progression rather than absolute baseline level. MoCA is exempt from anchoring as it is decoded on its bounded calibrated scale; streams with no observations within the encoding window are anchored using the subject-specific estimate from the auxiliary prediction head.

#### Model dimensions

The GRU-D encoder has hidden dimension *d*_*h*_ = 40. Each stream router MLP has hidden dimension 32 and produces latent states **z**_*X*_ (0) ∈ ℝ^8^.The ODE base networks 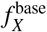 and drive networks 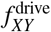 have hidden dimension 16. Readout heads are two-layer MLPs with hidden dimension 16. Overall, the student contains 39, 771 trainable parameters. The teacher shares the same downstream architecture and differs only in the encoder, employing a BiGRU-D with bidirectional access to the full training sequence to provide a temporally enriched supervisory signal for distillation.

#### Dropout regularization

Dropout is applied to the stream-specific routing networks and prediction heads with rates of 0.1 and 0.15, respectively. For uncertainty estimation, these dropout layers remain active during inference to perform Monte Carlo (MC) Dropout.

#### Training configuration

All models are implemented in PyTorch, with ODE integration performed using torchdiffeq and the adaptive Dormand–Prince (dopri5) solver, using a relative tolerance of 10^−7^ and an absolute tolerance of 10^−9^. Parameters are optimized with AdamW (Loshchilov and Hutter, 2017) (learning rate 3 × 10^−4^, weight decay 10^−3^), with gradients clipped to unit *ℓ*_2_-norm. A ReduceLROnPlateau scheduler halves the learning rate when validation MAE fails to improve for 10 consecutive epochs. Training uses batch size 32 for up to 200 epochs, with early stopping triggered after 30 epochs without validation improvement. Random seeds are fixed for data splitting, weight initialization, and dropout. All experiments are conducted on a single NVIDIA RTX A5000 (24 GB) GPU, with each training epoch requiring approximately 1 minute.

## 4. Experiments

We evaluate BEACON through ten complementary analyses spanning quantitative forecasting performance, biological interpretability, and clinical utility. ADNI is used for all experiments owing to its denser longitudinal trajectories and richer multimodal biomarker coverage (Figure 1); OASIS-3 is reserved for external validation.

### Evaluation metrics and statistical testing

Unless stated otherwise, all results report mean absolute error (MAE) of MoCA predictions over the extrapolation window, computed on the held-out test set. Statistical significance against BEACON is assessed throughout using the paired Wilcoxon signed-rank test on prediction errors.

### 4.1. Comparison with state-of-the-art methods

We benchmark BEACON against representative architectures spanning the principal paradigms for modelling irregular longitudinal clinical data: missingness-aware recurrent models (GRU-D (Che et al., 2018), T-LSTM (Baytas et al., 2017)); attention-based sequence models (Transformer (Vaswani et al., 2017), ContiFormer (Chen et al., 2023b)); hybrid recurrent-continuous-time models (ODE-RNN (Rubanova et al., 2019), GRUD-NODE, T-LSTM-NODE); a continuous-time neural differential equation model (Neural CDE-NODE); and a multimodal longitudinal AD progression model for incomplete variable-length trajectories (Xu et al., 2022).

While several AD-specific forecasting methods have been proposed recently (Bossa and Sahli, 2023; Zanin et al., 2025), they predominantly adopt variants of the same underlying temporal modeling backbones, differing primarily in feature engineering or auxiliary objectives rather than in the core sequence modeling framework. As publicly available implementations are unavailable for several of these methods, we benchmark against the canonical architectures foundational to these methods under a unified experimental protocol, enabling controlled comparison of temporal modeling capability independent of dataset-specific engineering choices.

To additionally isolate the contribution of the proposed biological priors, we evaluate unconstrained variants of BEACON in the component ablation (Section 4.5) that retain the same encoder and Neural ODE while removing the ATVNC cascade and monotonicity constraints.

### 4.2. Cross-Dataset Generalization

We assess cohort-level robustness of BEACON through two complementary transfer settings between ADNI and OASIS-3. In **zero-shot transfer**, the ADNI-trained model is evaluated directly on the OASIS-3 test set without adaptation, testing generalization across differing acquisition protocols and patient characteristics. In **fine-tuning**, the ADNI-pretrained model is further trained on the OASIS-3 training set before evaluation, assessing the efficiency of knowledge transfer from a large source cohort to a smaller target cohort.

### 4.3. Clinical Subgroup Analysis

To evaluate generalization across heterogeneous disease trajectories, we stratify the held-out test cohort into four clinically meaningful progression subgroups defined by diagnostic status derived from MoCA assessment at the start and end of the extrapolation window: stable cognitively normal (CN →CN), cognitively normal to MCI or dementia (CN → MCI/D), stable MCI (MCI →MCI), and progressive MCI or dementia (MCI → D, D →D). Within each subgroup, BEACON is compared against three SOTA baselines (GRUD-NODE, T-LSTM-NODE, and ODE-RNN) using extrapolation window MoCA prediction MAE. We further report densely predicted mean MoCA trajectories with 95% confidence intervals alongside the corresponding ground-truth mean trajectories for qualitative assessment.

### 4.4. Horizon-Stratified Evaluation

To characterize performance as a function of forecast horizon, we stratify extrapolation-window MoCA predictions into six temporal bands: 0-12, 12-24, 24-36,

36-48, 48-60 and 60 months beyond *T*_obs_. MAE are reported within each band for BEACON and the state-of-the-art baselines, to quantify where the biological constraints and teacher-student supervision provide the greatest benefit relative to unconstrained alternatives.

### 4.5. Ablation study I: Effect of BEACON components

We validate each architectural component through the following controlled variants, each obtained by removing or replacing a single design element from the full BEACON model (**A1**) (i) Without monotonicity (**A2**): The softplus output wrappers and directionality signs *ν*_*X*_ are removed from Eq. (9), allowing unrestricted non-monotone derivatives. A2 tests whether direction-aware constraints reduce trajectory violations and improve extrapolation; (ii) Without stream specific initialization (**A3**): The stream-specific routing networks *R*_*X*_ are removed; all streams are initialized from the shared encoder output, 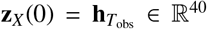. A3 tests the value of biologically partitioned latent initial conditions; (iii) Without hierarchical structure (**A4**): The inter-stream drive terms 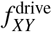 are removed, so each latent stream evolves independently. Although all streams are jointly initialized from observed clinical variables, no cross-stream interactions occur during latent evolution. A4 isolates the contribution of the ATVNC cascade; (iv) with shared NODE (**A5**): Stream-specific NODEs are replaced by a single shared NODE that jointly evolves all biomarker latents through a common readout head. A5 evaluates the benefit of stream-specific latent dynamics; finally, (v) without distillation (**A6**): The student model is trained end-to-end using ground-truth observations only, without teacher supervision (ℒ_con_ and ℒ_ali_ removed). A6 isolates the benefit of privileged teacher guidance.

### Biological plausibility

For each variant, we additionally report the *trajectory violation rate* (%), defined as the percentage of test subjects exhibiting at least one monotonicity reversal across the six constrained biomarker streams. To avoid penalizing measurement noise, reversals are counted only when the deviation from the expected monotonic trend exceeds 2% of the training-set standard deviation of the respective biomarker.

### 4.6. Ablation study II: Stream contributions

To quantify the contribution of individual pathological streams to MoCA forecasting, we train reduced variants in which the cognition NODE is coupled to only a single upstream stream: A+C, T+C, V+C, and N+C. These are compared against a cognition-only baseline (C) and the full ATVNC model, assessing the relative predictive importance of each biological pathway and if jointly modeling complementary disease mechanisms provides additive benefit.

### 4.7. Ablation study III: Loss components

We incrementally introduce the terms of the composite objective in Eq. (13) following the sequence (ℒ_ext_ +ℒ _rec_) → +ℒ _aux_ →+(ℒ_con_ + ℒ_ali_), to verify that each term provides complementary supervision and to quantify its individual contribution to long-horizon MoCA forecasting.

### 4.8. Biomarker trajectory fidelity

Although BEACON is primarily optimized for cognitive forecasting, we additionally assess the fidelity of the learned biomarker trajectories as a proxy for the biological plausibility of the latent ODE dynamics. Agreement between predicted and observed biomarker trajectories over the extrapolation window is evaluated using Lin’s concordance correlation coefficient (CCC) (Lawrence and Lin, 1989), reported per stream across all test subjects with at least one follow-up observation for that stream.

### 4.9. Clinical trial enrichment study

We evaluate whether BEACON forecasts can support prognostic enrichment for AD clinical trials by preferentially identifying participants at highest risk of clinical progression. For each test participant, risk is quantified as the negative predicted MoCA at a fixed future horizon (lower forecast MoCA = higher risk). Participants are ranked by this score, and the enrichment factor is computed as the conversion rate within the top-ranked subset divided by the cohort base rate. Two clinically relevant transitions are evaluated: MCI → D and CN →MCI/D at 24, 36, 48, and 60 months beyond *T*_obs_, at a primary enrollment depth of 35%, approximating a realistic screening yield while retaining sufficient conversion events for stable estimation.

### 4.10. Sensitivity and uncertainty analyses

#### Observation window sensitivity

Although BEA-CON is trained with a 24-month observation window, at inference we progressively restrict the available patient history to 6, 12, 18, and 24 months without retraining and evaluate extrapolation MAE at each horizon, assessing robustness to limited longitudinal data. This analysis also validates the choice of the 24-month window relative to shorter alternatives. **Uncertainty quantification**. Epistemic uncertainty is estimated using MC Dropout (Gal and Ghahramani, 2016) with dropout enabled at inference across 20 stochastic forward passes. Predictive mean and variance are computed per MoCA forecast, with the variance serving as the uncertainty estimate. Test subjects are ranked by predictive uncertainty and partitioned into ten equally sized deciles; MAE is reported within each decile to assess whether uncertainty estimates are calibrated with respect to forecasting reliability.

## 5. Results

### 5.1. Comparison with state-of-the-art methods

Table 2 presents MoCA forecasting performance on ADNI and OASIS-3 datasets against existing approaches. BEACON achieves the lowest overall MAE on ADNI (2.06 ± 0.08), significantly outperforming all competing methods (*p* < 0.05, paired Wilcoxon signed-rank test). Recurrent encoders specifically designed for irregular sampled, incomplete longitudinal clinical time series data (GRU-D and T-LSTM) and continuous time transformer architecture achieve stronger performance. However, hybrid architectures that retain recurrent encoding while employing Neural ODEs for latent evolution (T-LSTM-NODE and GRUD-NODE) further improve forecasting accuracy, with GRUD-NODE providing the strongest baseline performance (MAE = 2.19). Interestingly, replacing the recurrent encoders with continuous-time encoder (NCDE) degrades forecasting accuracy, suggesting that learning continuous-time representations directly from the limited 24-month observation window with sparse, irregularly sampled observations is challenging. BEACON, a hybrid architecture consistently surpasses all competing approaches, demonstrating that explicitly embedding biologically constrained latent disease dynamics provides complementary predictive information beyond advances in temporal sequence modeling alone.

The same trend is observed on OASIS-3, where BEACON achieves the best performance (MAE=1.90) despite being an independent cohort with differing acquisition characteristics and missing several ADNI biomarkers, including pTau, CMB counts, and DTI-derived measures. This cross-cohort consistency indicates that the learned biological priors generalize beyond the specific biomarker panel on which the model was trained.

### 5.2. Cross dataset generalization

Table 3 summarizes cross-cohort transfer performance. In the zero-shot setting, BEACON trained exclusively on ADNI achieves MAE=2.68 on OASIS-3, demonstrating generalization across cohorts with differing acquisition protocols and substantially different biomarker availability: OASIS-3 lacks pTau, the individually most predictive upstream ((Sperling et al., 2024), also evident from results in Section 5.6), as well as CMB counts and DTI-derived measures. The fact that the model achieves coherent forecasting under this degree of biomarker absence indicates that the biologically constrained latent dynamics do not depend on any single stream and degrade minimally under missing inputs. Fine-tuning on the OASIS-3 training cohort substantially reduces the MAE to 1.90, recovering full within-cohort performance and demonstrating effective knowledge transfer. While the 60-month bin in OASIS cohort contains only 14 visits, and its MAE is therefore based on a limited sample with a relatively large bootstrap standard error (0.68), BEACON maintains consistently strong performance across the remaining horizon bins and achieves the best overall extrapolation MAE in the OASIS cohort.

### 5.3. Clinical subgroup analysis

Figure 3(a) shows BEACON’s performance across the four clinical progression subgroups. The largest absolute advantage is observed in the two clinically most critical transition groups: CN to MCI or dementia (CN →MCI/D) and progressive MCI (MCI/D →D), precisely where accurate long-horizon forecasting has the greatest utility for trial enrichment and therapeutic decision-making. In both groups, BEACON closely tracks the observed longitudinal MoCA trajectory, whereas competing methods either systematically underestimate disease progression (GRU-D-NODE, ODE-RNN) or overpredict decline (T-LSTM-NODE), the latter leading to poor trajectory fidelity in the stable sub-groups despite competitive the overall MAE.

**Figure 3:**
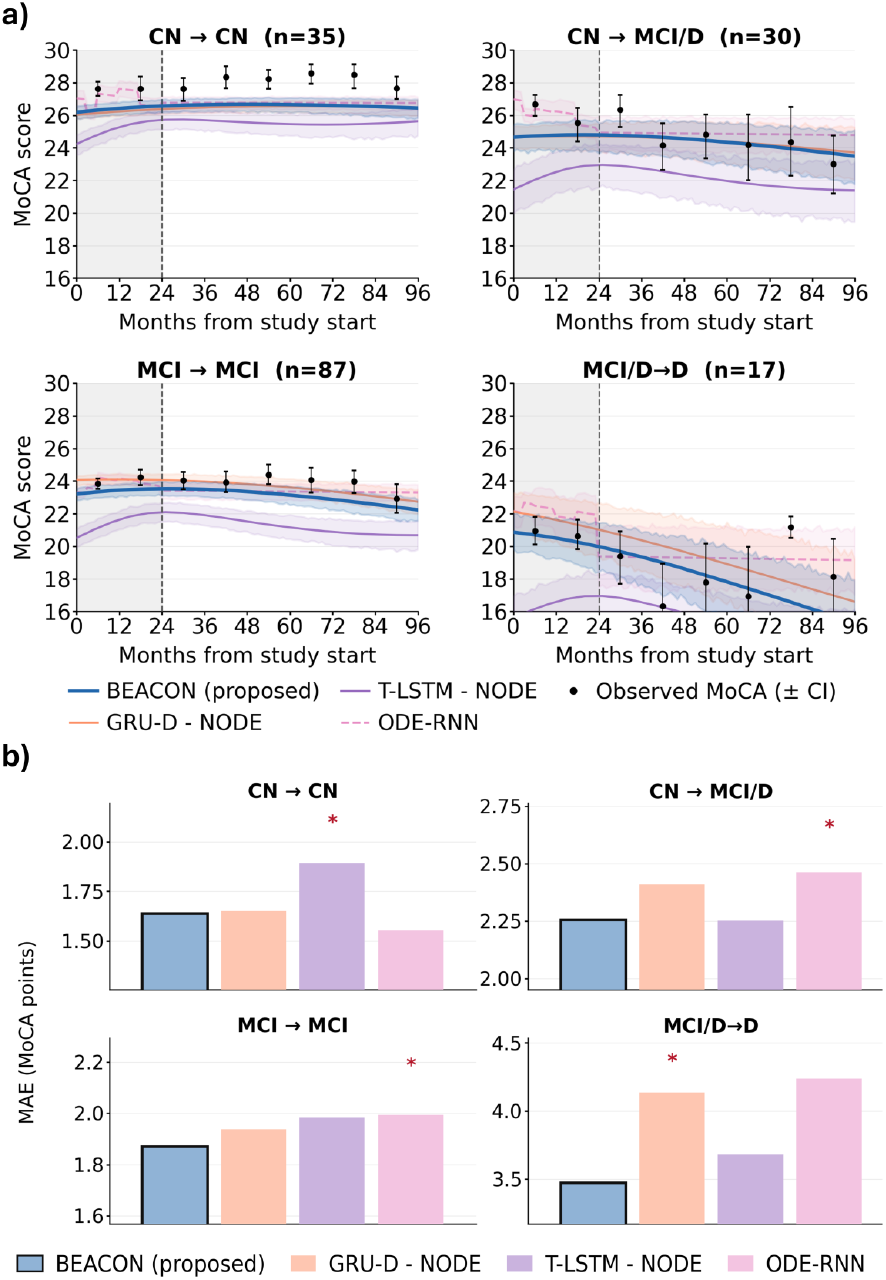
(a) Mean longitudinal MoCA trajectories with 95% boot-strap confidence intervals for BEACON and three SOTA baselines across four progression subgroups. (b) Quantitative comparison of extrapolation-window MoCA MAE per subgroup. Red asterisk indicates that the corresponding method performs significantly worse than BEACON in terms of MAE (p < 0.05).

Quantitatively, BEACON achieves the lowest error in MCI→MCI and MCI/D→D (Figure 3(b)), outperforming the strongest baseline by 3.4% (1.94 → 1.87) and 5.7% (3.68 → 3.47) respectively. In the CN→MCI/D subgroup, BEACON matches T-LSTM-NODE (2.26 vs. 2.25), however T-LSTM-NODE’s systematic overprediction (shown in Figure 3(a)) renders it unreliable across the remaining subgroups, limiting its clinical utility. Together, these results demonstrate that BEACON’s biologically grounded trajectory constraints produce more calibrated predictions across diverse clinical phenotypes than methods optimized for aggregate accuracy alone.

### 5.4. Horizon-Stratified Evaluation

BEACON consistently achieves the lowest MAE across all forecasting horizons in Table 2, with the performance gap widening at longer horizons where accurate extrapolation is most challenging. Relative to the strongest baseline (GRUD-NODE), BEACON reduces MAE by 8.7% at 36-48 months (2.42 →2.21) and by 8.2% beyond 60 months (2.43 → 2.23) on ADNI, while maintaining consistent improvements across all shorter intervals. Hybrid continuous-time models (ODE-RNN, GRUD-NODE, T-LSTM-NODE) exhibit improved long-range forecasting relative to discrete-time baselines, however BEACON’s biological constraints provide additional stabilization at the longest horizons. This is further corroborated by the component ablation (shown in Section 5.5, where removing monotonicity constraints consistently degrades performance at horizons beyond 36 months.

### 5.5. Ablation study I: Effect of BEACON components

Table 4 compares the full BEACON model (A1, MAE=2.06, 39,771 parameters) against five ablated variants. The complete model achieves the best MoCA forecasting accuracy at compact model size, demonstrating that biological structuring is both more effective and more parameter-efficient than its unconstrained alternatives. Eliminating the monotonicity constraints (A2) increases the MAE to 2.13, demonstrating that biologically constrained disease progression improves forecasting performance. Replacing stream-specific initializations with a shared encoder output (A3) more than doubles the parameter count (86,164) while increasing MAE to 2.12, confirming that biologically partitioned latent initial conditions yield a more compact and predictive representation. Removing the directed ATVNC cascade (A4) and stream-specific dynamics (A5) further degrades performance (MAE=2.14, 2.15), demonstrating that cross-stream hierarchical interactions provide complementary predictive information beyond independent stream evolution.

#### Distillation

Removing teacher supervision (A6) yields MAE=2.10 versus 2.06 for the full model. The modest absolute gain reflects the sparsity of available supervision: over 25% of participants in the train split have no MoCA observations and a further 25% have only a single assessment in the extrapolation window. Additionally, mean additional follow-up beyond the observation window is 1.20 visits for amyloid and 0.50 visits for pTau (Figure 1). Despite this, the teacher achieves substantially lower error (MAE=1.60) using complete trajectories, and the distillation consistently benefits long-horizon forecasting, most visibly beyond 60 months in the loss ablation (Section 5.7).

#### Biological plausibility

As shown in Figure 4(c), monotonicity-constrained variants (A1, A3, A4, A6) produce 0% trajectory violations, while the unconstrained variants (A2, A5) yield 35.4% and 28.1% violations respectively. Qualitatively, as shown in Figure 4(d), the unconstrained models produce decreasing pTau and amyloid trajectories in the absence of amyloid-clearance treatment and increasing or oscillatory hippocampal volumes, none of which are biologically feasible. These violations indicate that without monotonicity constraints, models exploit spurious temporal correlations in the sparse longitudinal observations, sacrificing biological plausibility for short-term predictive fit.

**Figure 4:**
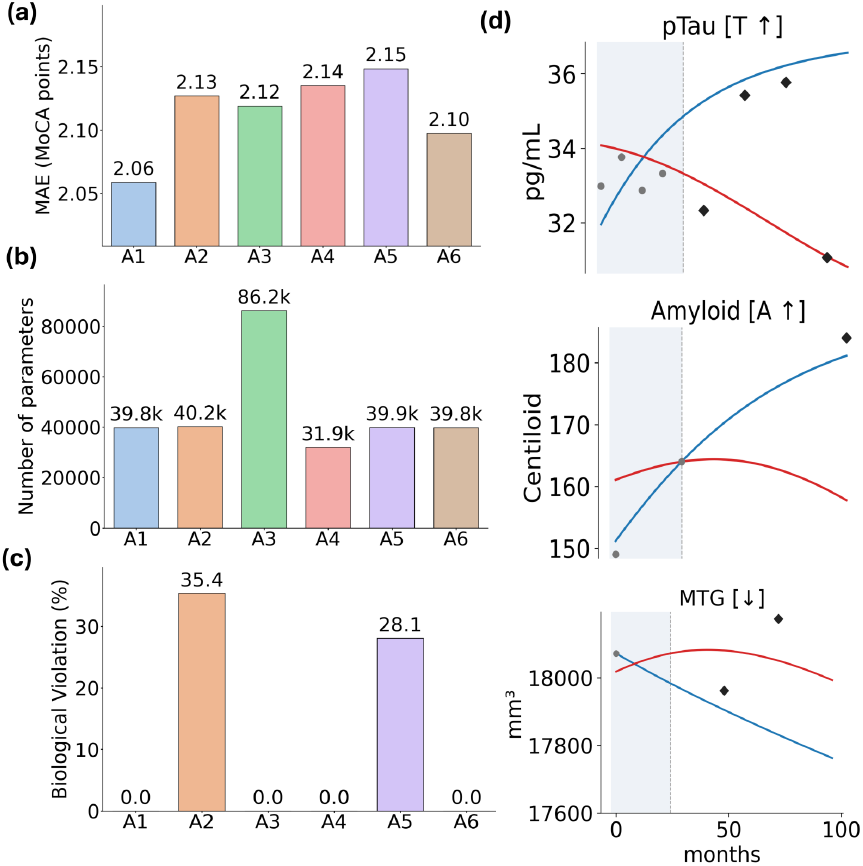
Component ablation study of the BEACON. Bar charts comparing A1-A6 across (a) MoCA extrapolation MAE, (b) total trainable parameters, and (c) biological violation rates (%). (d) Qualitative comparison of sample biomarker trajectories from the full BEACON model (blue) versus unconstrained variants A2 and A5 (red).Gray • and black _?_ indicate the true observations in the window (represented by dotted line) and extrapolation region respectively. MTG refers to Middle temporal gyrus volume.

### 5.6. Ablation study II: Stream contributions

Figure 5 shows MoCA forecasting MAE for the cognition-only baseline, single-stream variants, and the full ATVNC model. Incorporating upstream molecular pathology substantially improves prediction over cognitive history alone across all horizons. At short horizons, amyloid (A+C) and tau (T+C) yield the largest individual gains, reducing MAE by 8.6% and 8.0% respectively. While amyloid’s relative contribution diminishes at longer horizons (2.5%), consistent with early saturation of amyloid burden, tau retains the strongest long-horizon improvement (4.2%), reflecting its sustained propagation and close mechanistic association with cognitive decline. Vascular (V+C) and neurodegeneration (N+C) streams provide modest short-horizon gains but offer limited independent long-horizon information (≈ 1.5% gain), suggesting they primarily capture current disease state rather than encoding independent predictive signal for distant forecasting. The full ATVNC model consistently achieves the lowest MAE, with improvements of 6.9%, 9.7%, and 8.2% over the cognition-only baseline across short, medium, and long horizons respectively, demonstrating that jointly modeling interacting pathological pathways provides additive predictive benefit.

**Figure 5:**
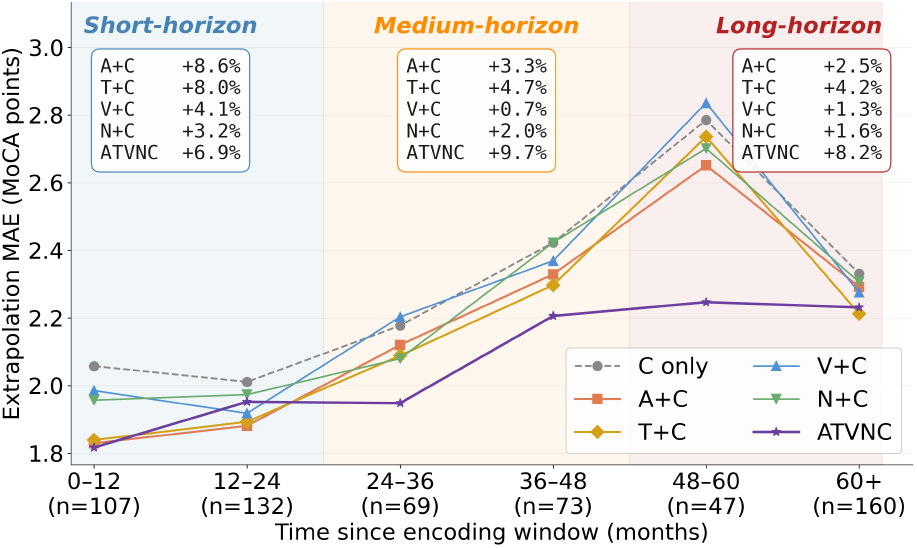
MoCA prediction MAE across forecasting horizons for the cognition-only, single-stream, and full ATVNC models. Short, medium, and long horizons denote 0–24, 24–48, and≥ 48 months, respectively.

### 5.7. Ablation study III: Loss components

Figure 6 shows the progressive effect of adding each loss component. Training with reconstruction loss alone (ℒ_ext_ + ℒ _rec_) yields the weakest performance, with prediction error increasing steadily with horizon. Adding ℒ_aux_ consistently improves accuracy across all horizons, reducing MAE from 1.75 to 1.68 at 36 months and from 2.37 to 2.23 at 72 months, confirming that supervising the latent initial conditions to reflect each subject’s stream-specific baseline disease burden produces more informative representations for downstream trajectory evolution. Further adding the distillation objective (ℒ _ali_ + ℒ _con_) provides additional improvement in long-horizon (beyond 60 months), with MAE decreasing from 2.23 to 2.15 at 72 months; the improvement is modest at short horizons for the reasons described in Section 5.5, but becomes progressively more pronounced beyond 60 months, where dense trajectory supervision from the teacher regularizes the latent ODE dynamics against observation sparsity. Together, the auxiliary initialization and distillation objectives provide complementary supervision, yielding more accurate and stable long-horizon forecasts than reconstruction loss alone.

**Figure 6:**
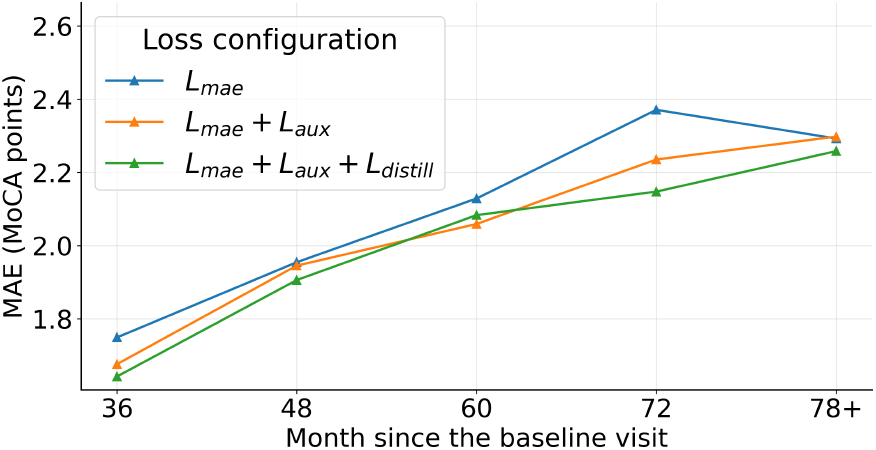
Loss ablation. MoCA MAE across forecasting horizons for additive loss configurations.

### 5.8. Biomarker trajectory fidelity

Despite being optimized primarily for cognitive forecasting, predicted biomarker trajectories remain well-aligned with observed longitudinal measurements. BEACON achieves CCC values between 0.75 and 0.89 across all streams (Figure 7), with the highest agreement for pTau, middle temporal gyrus, and CSF compartment. The comparatively lower CCC for WMH likely reflects the greater inter-individual heterogeneity of vascular pathology. These results confirm that the biologically constrained latent dynamics capture meaningful disease progression signals, validating the ATVNC representation as a coherent biological model of AD pathophysiology while primarily optimizing long-term cognitive forecasting.

**Figure 7:**
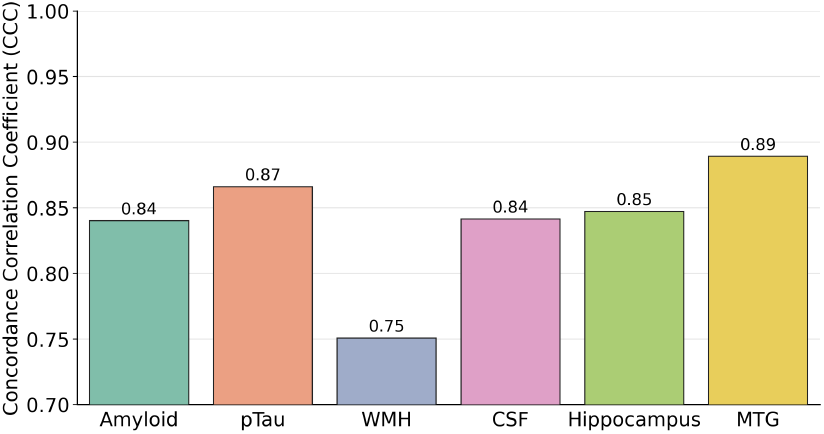
Lin’s concordance correlation coefficient (CCC) between the BEACON predicted and observed biomarker trajectories over the extrapolation window, reported per stream.

### 5.9. Clinical trial enrichment study

At the 35% enrollment depth, all methods concentrate future converters substantially above chance, achieving approximately 2.1–2.7 × enrichment over the cohort base rate across both diagnostic transitions and all horizons. BEACON achieves the highest or near-highest enrichment in all categories, reaching 2.7 × for MCI →D at 24 and 36 months, with consistent advantages for CN →MCI/D at 36–60 months. Notably, BEACON achieves this competitive enrichment despite substantial biomarker sparsity (∼ 69–70% missing amyloid and tau, ∼ 22–36% missing vascular, neurodegeneration, and cognitive markers) within the observation window, indicating that the biologically constrained ATVNC latent dynamics can infer coherent disease trajectories from incomplete multimodal observations and support reliable long-horizon prognostic ranking.

### 5.10. Sensitivity and uncertainty analysis

#### Observation window sensitivity

Figure 9(a) shows that BEACON is robust to substantial reductions in available clinical history. Shortening the encoding window from 24 months to 6 months increases the 60-month prediction MAE by only 0.21 (2.15 → 2.36), despite a 75% reduction in the observation period, a compression that reduces mean available visits from 3.5 to 1.7 for cognition and from 1.7 to 0.9 for amyloid (Figure 9(b)). Prediction curves remain closely aligned across all encoding durations, with the additional longitudinal information in the 24-month window primarily benefiting the most distant forecasts, reducing MAE from 2.03 to 1.78 at 78 months. These results validate the 24-month window choice, while demonstrating that biologically constrained latent dynamics enable stable forecasting even from minimal early clinical data.

#### Uncertainty quantification

MC Dropout uncertainty estimates are well-calibrated with respect to forecasting reliability. As subjects are ranked from the lowest to highest uncertainty decile, MAE increases monotonically from 1.65 to 2.89 MoCA points, yielding a strong positive correlation between uncertainty and error (Spearman *ρ* = 0.84, Figure 10). This indicates that the uncertainty estimates effectively identify predictions with lower confidence, providing a clinically useful signal for flagging cases that may benefit from additional assessment or more frequent monitoring and assist in interpreting the reliability of long-term cognitive predictions.

## 6. Discussion

In this study, we have proposed BEACON, a biologically constrained dual-encoder framework for long-horizon MoCA forecasting from sparse, irregularly sampled noisy multimodal longitudinal data, with missing biomarker modalities. On ADNI, BEACON achieves the lowest extrapolation MAE across all prediction horizons relative to nine competing baselines, with the performance advantage widening at horizons beyond 36 months where maintaining biologically coherent trajectories is most critical. Strong cross-cohort generalization to the independent OASIS-3 cohort, despite the distribution shift in recruitment, imaging protocols, processing pipelines and complete absence of pTau, DTI, and CMB modalities during evaluation, suggests that the learned latent representations encode cohort-agnostic biological features rather than site-specific or modality-dependent patterns.

An important consideration in evaluating long-horizon cognitive forecasting models is the relationship between statistical accuracy and clinical utility. The absolute MAE of 2.06 points on a 30-point scale is at the boundary of the minimal clinically important difference for MoCA (estimated at approximately 1-2.0 points (Lindvall et al., 2024)), and the improvement over the strongest baseline (0.13 points) falls below individual-level clinical discriminability. Therefore, rather than merely a tool for predicting individual patients’ MoCA scores with sufficient precision to guide treatment decisions at the bedside, BEACON’s clinical value also lies in three complementary domains.

Firstly, *cohort-level enrichment for clinical trials*: reliable trajectory models enable identification of fast progressors. This enrichment potential is supported by the subgroup trajectory analysis (Figure 3), where BEACON remains most closely aligned with observed long-horizon MoCA trajectories across clinical progression regimes. Consistent with this, the clinical trial enrichment analysis (Figure 8) shows that selecting the top 35% highest-risk participants concentrates future converters within the enrolled subset, yielding approximately 2.1-2.7 fold enrichment for both CN → MCI/D and MCI → D across the 24–60 month prediction horizons. Secondly, *direction-of-change estimation*: the clinically relevant question in longitudinal monitoring is frequently whether a patient is declining and at what rate, rather than the absolute score at a future time point. BEACON’s largest gains are observed in the MCI →D subgroup (5.7% MAE reduction as shown in Figure 3), precisely the population where trajectory direction is most consequential for therapeutic decision-making. Finally, and uniquely to our framework, *biological plausibility enforcement*: unconstrained baselines produce trajectory reversals (e.g., decreasing amyloid burden, recovering hippocampal volume) in up to 35% of test subjects without any disease-modifying intervention as seen in Figure 4. Such predictions are not only inaccurate, but also clinically misleading and could delay appropriate intervention if integrated into decision-support workflows. Unlike the compared recurrent networks and attention mechanisms, BEACON avoids such violations by design, representing a patient safety property that is independent of point-prediction accuracy.

**Figure 8:**
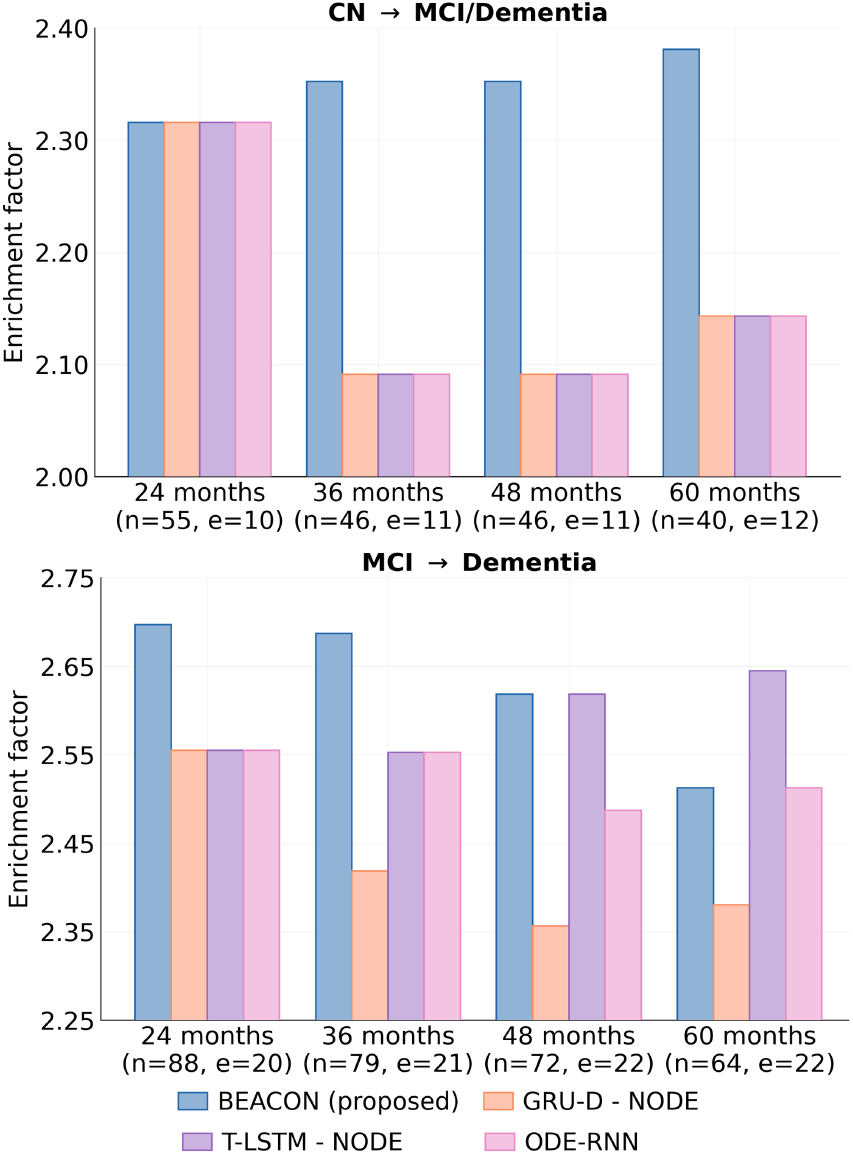
Clinical trial enrichment analysis. Enrichment factor (conversion rate in the top 35% highest-risk participants divided by the cohort base rate) for CN →MCI/dementia (top) and MCI → dementia (bottom) at 24, 36, 48, and 60 months beyond the observation window. Values above 1.0 indicate enrichment over chance. *n* = participants with available follow-up; *e* = converters.

**Figure 9:**
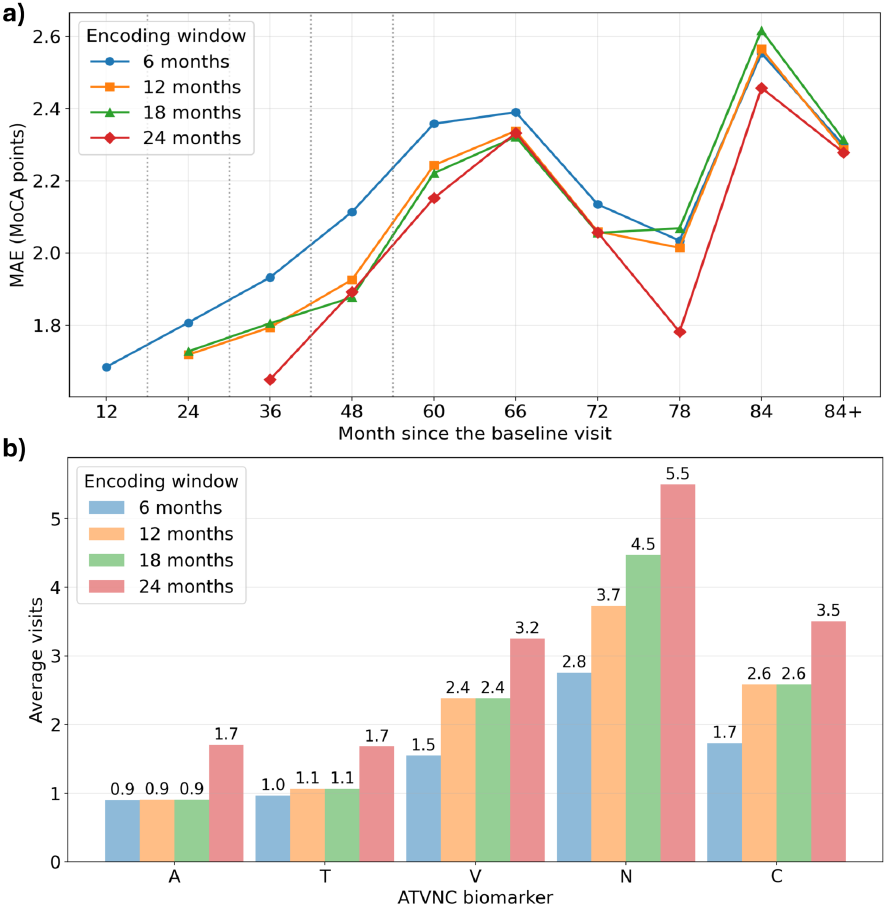
Observation window sensitivity. (a) MoCA MAE across prediction horizons for encoding windows of 6-24 months. (b) Mean available observations per biomarker stream within each window duration.

**Figure 10:**
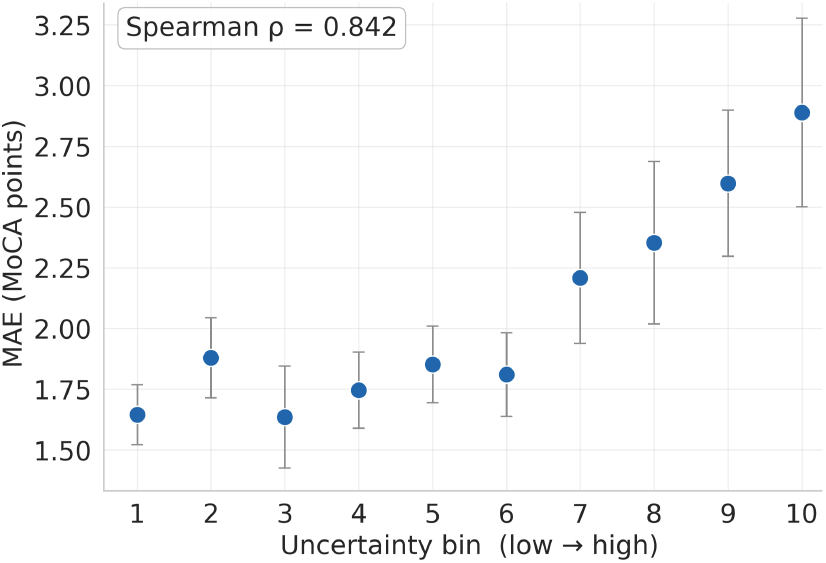
Forecasting reliability across uncertainty deciles. MAE is reported for predictions grouped into ten ordered uncertainty deciles (lowest to highest MC Dropout variance). Higher uncertainty deciles exhibit progressively larger forecasting errors.

Stream-specific latent initialization, conditioned on biologically relevant static covariates and supervised by per-stream auxiliary heads, provides a more precise representation of each participant’s baseline disease burden than a shared global latent state. The ablation demonstrates that this partitioned initialization is simultaneously more parameter-efficient (39k versus 86k parameters for the shared alternative as shown in Figure 4) and more predictively accurate, suggesting that biological disjointness in the latent space directly benefits downstream trajectory forecasting (Figures 4 and 6. The sensitivity analysis further demonstrates stable forecasting performance with as little as 6 months of baseline history (Figure 9), a practically important property given the high variability in clinical encounter frequency across care settings.

The stream ablation analysis in Figure 5 reveals a clinically interpretable pattern of predictive contributions: amyloid and tau provide the largest individual gains at short horizons (8.6% and 8.0% respectively), consistent with their upstream role as the primary molecular drivers of AD progression, while tau retains the strongest long-horizon advantage (5.1%), reflecting its sustained propagation and close mechanistic association with neurodegeneration and cognitive decline (Hardy and Higgins, 1992; Raj et al., 2025). The full ATVNC model consistently outperforms any single-stream variant, demonstrating that the complementary information encoded across pathological pathways is additive rather than redundant. Importantly, despite being optimized primarily for cognitive forecasting, BEACON reconstructs longitudinal biomarker trajectories with CCC of 0.75-0.89 as shown in Figure 7, confirming that the learned latent dynamics capture biologically meaningful disease evolution rather than merely fitting cognitive outcomes.

The benefit of teacher supervision is most evident at the longest forecast horizons: reducing the 72-month MAE from 2.23 to 2.15, consistent with the mechanism of dense trajectory distillation (Figure 6), whereby the teacher regularizes the latent ODE dynamics against observation sparsity throughout the extrapolation window, an effect that compounds where ground-truth supervision is most scarce. The modest overall distillation gain reflects a structural limitation of ADNI’s follow-up density: over 25% of participants lack MoCA observations in the extrapolation window, constraining the student’s available learning signal regardless of teacher quality.

Although BEACON demonstrates robust performance across heterogeneous disease stages, a few limitations remain. The ATVNC framework is tailored to AD pathophysiology and hence extension to other neurodegenerative disorders, including Lewy body disease and frontotemporal dementia, will require incorporating disease-specific biomarkers and biological priors. Although BEACON is robust to missing biomarker modalities, the current evaluation is restricted to the fixed biomarker panel available in ADNI and OASIS-3; integration of emerging plasma biomarkers (p-tau217, GFAP), digital cognitive assessments, and functional imaging may further improve predictive performance and clinical reach. BEACON does not model disease-modifying therapeutic effects, as such information is largely unavailable in these cohorts. Treatment-aware trajectory modeling is an important clinical extension, particularly given recent regulatory approvals of amyloid-targeting therapies (Van Dyck et al., 2023). The current framework also models cognitive decline as a single scalar outcome (MoCA); future work may extend this to domain-specific cognitive profiles capturing differential decline across memory, executive function, and language, which would provide finer-grained insight into the neuropsychological signature of individual disease trajectories. A further limitation is the scarcity of long-horizon follow-up data for advanced AD participants in both cohorts (Table 1) (most of AD cases contribute only less than 3 longitudinal visits), constraining the reliability of performance estimates at the longest forecast horizons in this subgroup. Finally, while subgroup analyses demonstrate meaningful performance differences across progression stages, prospective clinical validation, including evaluation of enrichment efficiency in a simulated trial design, assessment of threshold-crossing accuracy for MoCA clinical category transitions (cognitively normal, MCI, dementia), and calibration of predictive uncertainty against observed outcomes, is necessary before deployment in any decision-support context.

**Table 1:** Summary statistics of key clinical, cognitive, genetic, and neuroimaging variables for the OASIS and ADNI cohorts. APOE genotype is reported as allele pairs (*ϵ*2/*ϵ*2–*ϵ*4/*ϵ*4), where *ϵ*2 is generally protective and ϵ4 confers progressively greater genetic risk for Alzheimer’s disease. APOE genotype data is unavailable for 5 participants in the OASIS cohort overall. Asterisks in the OASIS columns indicate subsets with missing APOE data (^*^=one participant; ^**^=two participants).

| Split | Diagnosis | N | Age | MoCA | Amyloid | pTau | APOE Allele Status N |  |  |  |  |  |
| --- | --- | --- | --- | --- | --- | --- | --- | --- | --- | --- | --- | --- |
|  |  |  | (years) | (0–30) | (Centiloid) | (pg/mL) | 2/2 | 2/3 | 3/3 | 2/4 | 3/4 | 4/4 |
| ADNI Dataset |  |  |  |  |  |  |  |  |  |  |  |  |
| Train | CN | 399 | 72.0 ± 6.2 | 25.8 ± 2.6 | 19.2 ± 33.8 | 21.1 ± 9.5 | 0 | 39 | 237 | 7 | 102 | 14 |
|  | MCI | 427 | 71.8 ± 7.5 | 22.8 ± 3.4 | 43.3 ± 48.9 | 27.1 ± 14.5 | 0 | 22 | 196 | 5 | 155 | 49 |
|  | AD | 129 | 73.9 ± 8.1 | 17.5 ± 4.3 | 83.1 ± 45.0 | 36.7 ± 16.1 | 1 | 5 | 37 | 3 | 57 | 26 |
| Val | CN | 33 | 72.9 ± 6.1 | 25.7 ± 2.8 | 9.5 ± 30.4 | 20.0 ± 8.7 | 0 | 3 | 19 | 0 | 9 | 2 |
|  | MCI | 54 | 71.3 ± 6.8 | 23.8 ± 2.9 | 37.0 ± 46.7 | 26.9 ± 17.2 | 0 | 1 | 30 | 0 | 15 | 8 |
|  | AD | 5 | 81.8 ± 6.3 | 18.6 ± 3.1 | 44.4 ± 59.9 | 37.7 ± 15.5 | 0 | 0 | 1 | 0 | 4 | 0 |
| Test | CN | 72 | 72.7 ± 5.8 | 25.7 ± 2.7 | 29.2 ± 46.3 | 23.5 ± 8.8 | 0 | 12 | 38 | 1 | 21 | 0 |
|  | MCI | 108 | 71.5 ± 7.3 | 24.0 ± 3.0 | 36.4 ± 45.3 | 25.0 ± 13.8 | 0 | 11 | 51 | 2 | 38 | 6 |
|  | AD | 8 | 71.6 ± 10.1 | 17.4 ± 4.9 | 86.8 ± 48.1 | 36.4 ± 16.9 | 0 | 0 | 2 | 0 | 3 | 3 |
| OASIS Dataset |  |  |  |  |  |  |  |  |  |  |  |  |
| Train | CN | 113 | 66.4 ± 8.2 | 28.1 ± 1.1 | 8.8 ± 15.1 | - | 0 | 12 | 60 | 10 | 25 | 6 |
|  | MCI * | 84 | 68.1 ± 6.9 | 25.1 ± 0.8 | 10.8 ± 21.4 | - | 1 | 8 | 38 | 2 | 30 | 4 |
|  | AD | 52 | 72.0 ± 6.9 | 20.3 ± 2.7 | 35.9 ± 36.4 | - | 0 | 7 | 19 | 2 | 20 | 4 |
| Val | CN * | 43 | 64.2 ± 8.4 | 28.1 ± 1.1 | 5.1 ± 8.0 | - | 1 | 4 | 24 | 1 | 12 | 0 |
|  | MCI | 32 | 67.3 ± 7.5 | 25.2 ± 0.8 | 19.8 ± 28.9 | - | 0 | 2 | 23 | 0 | 6 | 1 |
|  | AD | 25 | 72.2 ± 9.5 | 20.2 ± 3.5 | 19.8 ± 26.2 | - | 0 | 3 | 9 | 1 | 9 | 3 |
| Test | CN ** | 116 | 66.8 ± 7.4 | 28.0 ± 1.0 | 8.5 ± 16.1 | - | 0 | 12 | 66 | 5 | 26 | 5 |
|  | MCI * | 102 | 68.1 ± 7.3 | 25.1 ± 0.8 | 8.0 ± 14.1 | - | 1 | 17 | 53 | 3 | 22 | 5 |
|  | AD | 82 | 72.8 ± 7.5 | 19.9 ± 3.1 | 16.0 ± 24.3 | - | 1 | 8 | 40 | 2 | 28 | 3 |

**Table 2:** Forecasting performance across temporal prediction bins. MAE and bootstrap standard error (500 resamples) are reported over the overall extrapolation window and within each temporal bin for ADNI and OASIS-3. Column headers indicate prediction horizon in months; *n* denotes the number of visits per bin (ADNI/OASIS-3). Best-performing method is highlighted in bold and asterisks denote statistically significant differences relative to BEACON (^∗^ *p* < 0.05, ^∗∗^ *p* < 0.01, ^∗∗∗^ *p* < 0.001, paired Wilcoxon signed-rank test).

| SOTA Method | Overall MAE | Binned MAE |  |  |  |  |  |
| --- | --- | --- | --- | --- | --- | --- | --- |
|  |  | 0–12<br>(n=107/101) | 12–24<br>(n=132/203) | 24–36<br>(n=69/164) | 36–48<br>(n=73/116) | 48–60<br>(n=47/74) | >60<br>(n=160/14) |
| ADNI |  |  |  |  |  |  |  |
| Recurrent models |  |  |  |  |  |  |  |
| GRU-D | 2.25(0.08)<sup>***</sup> | 2.02(0.15) | 2.16(0.15) | 2.09(0.24) | 2.38(0.23) | 2.67(0.34) | 2.35(0.18) |
| T-LSTM | 2.27(0.08)<sup>***</sup> | 1.96(0.15) | 2.20(0.14) | 2.17(0.24) | 2.34(0.23) | 2.70(0.34) | 2.42(0.19) |
| Xu et al. (2022) | 2.66(0.12)<sup>***</sup> | 2.20(0.21) | 2.37(0.21) | 3.60(0.83) | 2.57(0.26) | 2.84(0.41) | 3.03(0.28) |
| Attention-based sequence models |  |  |  |  |  |  |  |
| Transformer | 2.27(0.08)<sup>***</sup> | 1.97(0.17) | 2.27(0.15) | 2.12(0.24) | 2.31(0.23) | 2.64(0.35) | 2.43(0.20) |
| ContiFormer | 2.23(0.08)<sup>*</sup> | 2.06(0.12) | 2.07(0.14) | 1.94(0.23) | 2.24(0.22) | 2.59(0.41) | 2.45(0.21) |
| Hybrid recurrent-continuous-time model |  |  |  |  |  |  |  |
| ODE-RNN | 2.22(0.09)<sup>*</sup> | 2.00(0.14) | 2.12(0.12) | 2.04(0.26) | 2.27(0.24) | 2.56(0.41) | 2.39(0.23) |
| T-LSTM-NODE | 2.19(0.08)<sup>*</sup> | 1.90(0.15) | 2.08(0.13) | 2.05(0.25) | 2.43(0.23) | 2.44(0.39) | 2.36(0.19) |
| GRUD-NODE | 2.19(0.08)<sup>*</sup> | 1.86(0.12) | 2.02(0.12) | 2.06(0.22) | 2.42(0.20) | 2.35(0.32) | 2.43(0.19) |
| Continuous time NODE models |  |  |  |  |  |  |  |
| NCDE - NODE | 2.35(0.09)<sup>***</sup> | 2.04(0.14) | 2.07(0.15) | 2.40(0.26) | 2.45(0.24) | 2.47(0.40) | 2.68(0.21) |
| BEACON (proposed) | **2.06(0.08)** | **1.82(0.12)** | **1.96(0.13)** | **1.95(0.22)** | **2.21(0.21)** | **2.24(0.31)** | **2.23(0.19)** |
| OASIS |  |  |  |  |  |  |  |
| Recurrent models |  |  |  |  |  |  |  |
| GRU-D | 2.03(0.06)<sup>***</sup> | 2.03(0.15) | 2.00(0.11) | 1.92(0.13) | 2.18(0.16) | 2.15(0.18) | 1.65(0.26) |
| T-LSTM | 2.06(0.06)<sup>**</sup> | 2.13(0.15) | 2.02(0.10) | 1.97(0.13) | 2.20(0.14) | 2.16(0.19) | 1.53(0.34) |
| Xu et al. (2022) | 4.53(0.17)<sup>***</sup> | 4.66(0.46) | 4.27(0.28) | 4.77(0.36) | 5.01(0.52) | 3.80(0.46) | 4.78(0.75) |
| Attention-based sequence models |  |  |  |  |  |  |  |
| Transformer | 2.13(0.07)<sup>*</sup> | 2.02(0.16) | 2.30(0.12) | 2.04(0.17) | 2.13(0.16) | 2.03(0.23) | 1.85(0.41) |
| ContiFormer | 2.03(0.06)<sup>*</sup> | 1.98(0.15) | 2.09(0.13) | 2.05(0.14) | 1.91(0.14) | 2.11(0.18) | 1.97(0.49) |
| Hybrid recurrent-continuous-time model |  |  |  |  |  |  |  |
| ODE-RNN | 2.00(0.06)<sup>*</sup> | 2.12(0.18) | 2.05(0.11) | 1.85(0.13) | 2.05(0.16) | 2.01(0.18) | 1.43(0.29) |
| T-LSTM-NODE | 2.04(0.06)<sup>**</sup> | 2.15(0.18) | 2.14(0.12) | 1.88(0.12) | 2.00(0.14) | 1.99(0.20) | 1.88(0.25) |
| GRUD-NODE | 2.01(0.06)<sup>*</sup> | 2.10(0.18) | 2.09(0.11) | 1.87(0.13) | 2.00(0.13) | 2.01(0.19) | 1.92(0.21) |
| Continuous time NODE models |  |  |  |  |  |  |  |
| NCDE-NODE | 2.15(0.06)<sup>***</sup> | 2.25(0.16) | 2.26(0.12) | 1.92(0.13) | 2.15(0.14) | 2.30(0.19) | 1.49(0.32) |
| BEACON | **1.90(0.06)** | **1.87(0.16)** | **1.89(0.12)** | **1.90(0.14)** | **1.95(0.14)** | **1.86(0.19)** | **2.18(0.68)** |

**Table 3:** Cross-dataset generalization. MAE and bootstrap standard error (500 resamples) across temporal bins (months). *n* denotes visit counts per bin (ADNI/OASIS-3).

| Training → Evaluation | Overall MAE | Binned MAE |  |  |  |  |  |
| --- | --- | --- | --- | --- | --- | --- | --- |
|  |  | 0–12<br>(n=107/101) | 12–24<br>(n=132/203) | 24–36<br>(n=69/164) | 36–48<br>(n=73/116) | 48–60<br>(n=47/74) | >60<br>(n=160/14) |
| ADNI → ADNI | 2.06(0.07) | 1.82(0.12) | 1.96(0.12) | 1.95(0.21) | 2.21(0.21) | 2.24(0.30) | 2.23(0.19) |
| ADNI → OASIS<br>(Zero-shot) | 2.68(0.09) | 2.62(0.23) | 2.50(0.15) | 2.68(0.18) | 2.72(0.23) | 3.05(2.98) | 3.53(0.80) |
| ADNI → OASIS<br>(Fine-tuned) | 1.90(0.06) | 1.87(0.16) | 1.89(0.12) | 1.90(0.14) | 1.95(0.14) | 1.86(0.19) | 2.18(0.68) |

## 7. Conclusions

We introduced BEACON, a biologically constrained dual-encoder framework that embeds the ATVNC disease cascade as a structural prior within continuous-time latent dynamics for long-horizon MoCA forecasting. Consistent outperformance of state-of-the-art baselines, elimination of biologically implausible trajectory violations, and strong generalization to an independent cohort collectively demonstrate that incorporating established disease knowledge as architectural priors within continuous-time generative models is a principled and effective approach to longitudinal neurodegenerative disease modeling. Future work will explore treatment-aware trajectory modeling, integration of emerging plasma biomarkers, and extension to domain-specific cognitive profiles and other neurodegenerative disorders.

## 8. Data availability and Ethics statement

This study used the de-identified data from the Alzheimer’s Disease Neuroimaging Initiative (ADNI; https://adni.loni.usc.edu/) and the Open Access Series of Imaging Studies (OASIS-3; https://www.nitrc.org/projects/oasis3/) databases. The original ADNI and OASIS-3 studies received approval from their respective Institutional Review Boards (IRBs), and written informed consent was obtained from all participants. Access to both datasets was obtained through the respective data access procedures and data use agreements administered by the ADNI and OASIS repositories. The present work involved secondary analysis of existing de-identified research data and did not involve direct interaction with human participants. Accordingly, no additional institutional ethical approval was required for this retrospective secondary analysis under the applicable data use agreements.

## Declaration of Generative AI and AI-assisted technologies in the writing process

This manuscript was written entirely by the authors without the use of large language models (LLMs) or artificial intelligence tools. All content, including analysis, interpretations, and conclusions, is solely the product of the authors’ research and intellectual effort.

## Acknowledgments

Data collection and sharing for the Alzheimer’s Disease Neuroimaging Initiative (ADNI) is funded by the National Institute on Aging (National Institutes of Health Grant U19AG024904). The grantee organization is the Northern California Institute for Research and Education. In the past, ADNI has also received funding from the National Institute of Biomedical Imaging and Bioengineering, the Canadian Institutes of Health Research, and private sector contributions through the Foundation for the National Institutes of Health (FNIH) including generous contributions from the following: AbbVie, Alzheimer’s Association; Alzheimer’s Drug Discovery Foundation; Araclon Biotech; BioClinica, Inc.; Biogen; Bristol Myers Squibb Company; CereSpir, Inc.; Cogstate; Eisai Inc.; Elan Pharmaceuticals, Inc.; Eli Lilly and Company; EuroImmun; F. Hoffmann-La Roche Ltd and its affiliated company Genentech, Inc.; Fujirebio; GE Healthcare; IXICO Ltd.; Janssen Alzheimer Immunotherapy Research & Development, LLC.; Johnson & Johnson Pharmaceutical Research & Development LLC.; Lumosity; Lundbeck; Merck & Co., Inc.; Meso Scale Diagnostics, LLC.; NeuroRx Research; Neurotrack Technologies; Novartis Pharmaceuticals Corporation; Pfizer Inc.; Piramal Imaging; Servier; Takeda Pharmaceutical Company; and Transition Therapeutics. For OASIS-3, Longitudinal Multimodal Neuroimaging: Principal Investigators: T. Benzinger, D. Marcus, J. Morris; NIH P30 AG066444, P50 AG00561, P30 NS09857781, P01 AG026276, P01 AG003991, R01 AG043434, UL1 TR000448, R01 EB009352. AV-45 doses were provided by Avid Radiopharmaceuticals, a wholly owned subsidiary of Eli Lilly.

